# Real-World Performance of the 2026 AHA/ACC Pulmonary Embolism Framework in a Multi-System CTPA Cohort

**DOI:** 10.64898/2026.08.06.26359865

**Authors:** Mahmoud Alwakeel, Suraj Zaveri, Emory Buck, Sudarshan Rajagopal, Divya Verma, Daniel Loriaux, Ricardo Henao, Victor F. Tapson, Thomas L. Ortel, William Schuyler Jones, Jon Martin, Krista L. Haines, Nikki L. B. Freeman, An-Kwok Ian Wong

## Abstract

**Background:** The 2026 American Heart Association/American College of Cardiology (AHA/ACC) guidelines replaced the 2019 European Society of Cardiology (ESC) four-tier pulmonary embolism (PE) risk scheme with five clinical categories (A–E) and subcategories. These categories were set by expert consensus and have not been validated against outcomes. How patients are reclassified relative to ESC, or how the two systems compare prognostically, is unknown.

**Methods:** We utilized three cohorts of patients with confirmed PE using structured electronic health record data, laboratory biomarkers, and large-language-model abstraction of radiology reports: Duke University Health System (n=12,992, drawn from 95,760 consecutive inpatient CT pulmonary angiography studies, 2014–2025, with no referral or registry enrollment step between imaging and cohort entry), INSPECT (Stanford; n=3,870), and MIMIC-IV (Beth Israel Deaconess; n=361). Patients were assigned AHA/ACC categories B through E, subcategorized where data allowed, and mapped to 2019 ESC risk strata. The primary outcome was 30-day mortality; discrimination was assessed with Harrell C-index.

**Results:** Among 17,223 patients with confirmed PE, pooled 30-day mortality rose monotonically across categories: 1.5% (B), 8.9% (C), 15.5% (D), and 31.9% (E), with the ordering preserved in all three cohorts despite differing baseline mortality. Subcategory-level discrimination was reliable only at the high-acuity extreme (D2–E2); across subcategories C1 through D1, mortality did not order monotonically (9.2%, 10.8%, 8.1%, 10.9%), and adding subcategories to category C did not improve discrimination at Duke (C-index 0.699 vs 0.699). Category C patients lacking both echocardiography and biomarker testing (12.7% of category C) had mortality (10.4%) equal to or exceeding classified peers. Relative to ESC, the frameworks were concordant at the extremes, but 5.7%of ESC intermediate-risk patients were reclassified to category D, with modestly higher but non-significant 30-day mortality than those remaining in category C (10.8% versus 8.9%).

**Conclusions:** Across a three-health-system cohort, the 2026 AHA/ACC framework produced a reproducible mortality gradient at the category level, with added subcategory granularity refining risk chiefly at the highest-acuity tiers. Discrimination across the broad intermediate band was limited, and reclassification from ESC fell almost entirely within this range.

## Background

Pulmonary embolism (PE) is the third most common cause of cardiovascular death and accounts for approximately 100,000 deaths annually in the United States.^1^ Several studies suggest that mortality has been *increasing* over the past several decades, in spite of recent advances in management.^2–7^ Its clinical course is markedly heterogeneous, ranging from patients who can be managed as outpatients on anticoagulation alone to those who deteriorate rapidly into obstructive shock and in-hospital death.^8,9^ This breadth is mirrored by equally broad therapeutic options, each with a distinct balance of benefit and risk.^10,11^ Since both under- and overtreatment carry harm, accurate risk stratification is central to PE management. Early prognostic tools such as the Pulmonary Embolism Severity Index (PESI, 2005) and its simplified form (sPESI, 2010) combined age, comorbidities, and vital signs to identify patients at low risk of 30-day mortality suitable for outpatient care.^12,13^ Subsequent frameworks incorporated PE-specific pathophysiology, such as evidence of right ventricular (RV) dysfunction on imaging and elevated cardiac biomarkers, alongside these scores.^14,15^ The 2019 European Society of Cardiology (ESC) guidelines integrated hemodynamic status, RV dysfunction, and biomarkers into a four-tier scheme of low, intermediate-low, intermediate-high, and high risk that became the prevailing standard in practice.^10^

The 2026 American Heart Association/American College of Cardiology (AHA/ACC) guideline departs substantially from this approach.^16^ In place of an ordinal low-to-high scale, it defines five clinical categories (A–E) with subcategories, assigning each patient by the most severe clinical, laboratory, or imaging finding present. The framework draws on the same domains as the ESC system—clinical severity scores (PESI, sPESI, or Bova), biomarkers, RV imaging, and hemodynamics—but adds granularity at both ends of the spectrum. At the low-acuity end, it separates patients sometimes suitable for discharge from those warranting brief observation, and at the high-acuity end, it subdivides what the ESC framework treated as a single high-risk tier, distinguishing incipient cardiopulmonary failure (Category D) from overt failure (Category E). A respiratory modifier (R) was also appended to any category to capture pulmonary compromise along a separate axis, although no recommendations were made for patients with this modifier. The categories are expert-consensus constructs that have not been validated against outcomes, and how patients map between the two systems, and how many are reclassified to higher or lower acuity, with the attendant implications for treatment escalation, has not been characterized.

Addressing these questions in routine practice has been constrained by the available evidence base, which derives largely from registries and referral populations that depend on labor-intensive manual abstraction that omit the broader denominator of patients evaluated for suspected PE.^17^ Additionally, the variables central to stratification, such as clot location and RV strain, reside in free-text radiology reports rather than structured fields. Recent advances in large language models (LLMs) now enable accurate, scalable extraction of these elements, making it feasible to assemble cohorts that represent the full population evaluated for suspected PE.^18,19^ Here, we assembled three cohorts representing academic medical centers across the US. Our largest is a system-wide cohort of consecutive inpatient computed tomography pulmonary angiography (CTPA) evaluations across the Duke University Health System, integrating structured electronic health record (EHR) data, biomarkers, imaging, and LLM-assisted abstraction of radiology reports, capturing both confirmed and excluded PE across academic and community hospitals. This is supplemented by INSPECT, representing Stanford University, (Palo Alto, CA) and MIMIC-IV, representing Beth Israel Deaconess (Boston, MA).^20,21^ Here, we quantified how the 2026 AHA/ACC categories reclassify patients relative to 2019 ESC stratification and compared the prognostic discrimination of the two systems for 30-day survival.

## Methods

### Cohort Definition and Dataset Composition

We included all adult (age ≥18) emergency department and inpatient encounters at the Duke University Health System (DUHS), comprising one academic teaching hospital, two community hospitals, and affiliated emergency departments and clinics, in which a CTPA was obtained for suspected pulmonary embolism between 2014 and 2025, regardless of final diagnosis. CTPA reports were abstracted using the peer-reviewed framework of Alwakeel et al for PE presence and location, right heart strain, and study quality;^22^ structured data, imaging and procedure reports, and death dates came from Epic Clarity (Epic Systems, Verona, WI), EKGs from MUSE (GE Healthcare, Chicago, IL), and comorbidities from ICD-10 codes using the Charlson index.^23^ Two external cohorts were assembled with the same abstraction framework, both deidentified and approved for use without informed consent. MIMIC-IV covers Beth Israel Deaconess Medical Center (Boston, MA) from 2008 to 2019, excluding CTPA not tied to an ICU admission, with reports identified from MIMIC-IV-NOTE and no capture of procedures such as suction thrombectomy.^22,24,25^ INSPECT covers Stanford University Hospital (Palo Alto, CA) from 2000 to 2021, excluding outpatient studies, with structured data reformatted from MEDS-OMOP to OMOP and no echocardiography or procedural data.^26,27^ The study was approved by the DUHS Institutional Review Board (Pro00115368).^28^

### Risk stratification

CTPA acquisition time was time zero. Vital signs and laboratory values were captured from 12 hours before to 24 hours after the first CTPA of the admission, and other diagnostic studies, including transthoracic echocardiography (TTE) and right heart catheterization (RHC), from 7 days before to 30 days after or until discharge, whichever came first. RV dysfunction on CT was defined by RV dilation, interventricular septal flattening or bowing, an RV/LV ratio above 0.9, or contrast reflux into the inferior vena cava on the CTPA report, in adherence with PERT consortium standardized data elements.^29^ Vasopressor and inotrope exposure came from medication administration records, and cardiac arrest from flowsheets. Patients were stratified independently by the 2026 AHA/ACC and 2019 ESC frameworks, each evaluated in descending severity.^30,31^ Since all patients underwent CTPA, none met AHA/ACC category A. Entry to AHA/ACC categories C through E used sPESI, PESI, and Bova; ESC intermediate strata used sPESI and PESI.^13,32,33^ Hestia was not calculated, as the cohort was inpatient-focused.^34^ Category C was subclassified by TTE and cardiac biomarkers (troponin and B-type natriuretic peptide); patients with neither obtained in the ascertainment window were designated C-indeterminate, a data-availability state rather than an AHA/ACC category.

### Statistical Analysis

Continuous variables are reported as means with standard deviations or medians with interquartile ranges according to distribution, categorical variables as counts and percentages, and analyses were stratified by PE status on the index CTPA. Group comparisons used chi-squared, Student’s t-test, or Mann-Whitney U tests; because a cohort of this size returns significant differences of limited clinical relevance, baseline comparisons are descriptive and p-values are not emphasized. Inferential analyses used p<0.05. Thirty-day survival was estimated by Kaplan-Meier, censoring patients alive at 30 days.^35^ Mortality proportions are reported with 95% Wilson score intervals and between-subcategory differences with 95% Newcombe hybrid intervals built from

Wilson limits (method 10).^36,37^ Cox proportional-hazards models used classification as the sole term with Efron handling of ties, so hazard ratios describe discrimination rather than an adjusted effect, and proportional hazards were tested with scaled Schoenfeld residuals.^38–40^ Within the intermediate band, a four-level model (C1, C2, C3, D1) was compared against a single pooled-band model by likelihood-ratio test, and ordered trend by Cochran-Armitage test and by a Cox model entering subcategory as one ordinal term, reported as a hazard ratio per step.^41,42^ Discrimination was quantified by the Harrell C-index for the full classification (B, C1, C2, C3, D1, D2, E1, E2) and for a collapsed classification pooling C1 through D1 (B, pooled band [C1-D1], D2, E1, E2), with their difference and its 95% confidence interval from a paired nonparametric bootstrap (500 resamples, 2.5th to 97.5th percentiles, the same resample applied to both).^43,44^ Equivalence within the band was tested by two one-sided tests against prespecified margins of 3.0 and 2.0 percentage points on the absolute risk-difference scale; within-band comparisons are exploratory and unadjusted for multiplicity.^45,46^ Analyses were complete-case with no imputation: where a required result was not obtained, the criterion was treated as not met, mirroring bedside application. Since an unordered test may itself carry prognostic information, this biases toward lower acuity, and its consequences are examined in the C-indeterminate sensitivity analysis.^47^ Missingness is reported as a cohort characteristic, for example the proportion without a cardiac troponin or without right-ventricular assessment. Analyses were performed in Python 3.12.^48,49^

## Results

### Cohorts and denominator

A total of 17,223 patients with confirmed PE were jointly studied across three academic health systems: 12,992 at Duke, 3,870 in INSPECT, and 361 in MIMIC-IV (**Figure 1**, **Table 1, Tables S1a-S1c**). Patients were distributed across AHA/ACC categories B-E: B: 4354 (25.3%), C: 9936 (57.7%), D: 1374 (8.0%), E: 1559 (9.1%).

**Figure 1.**
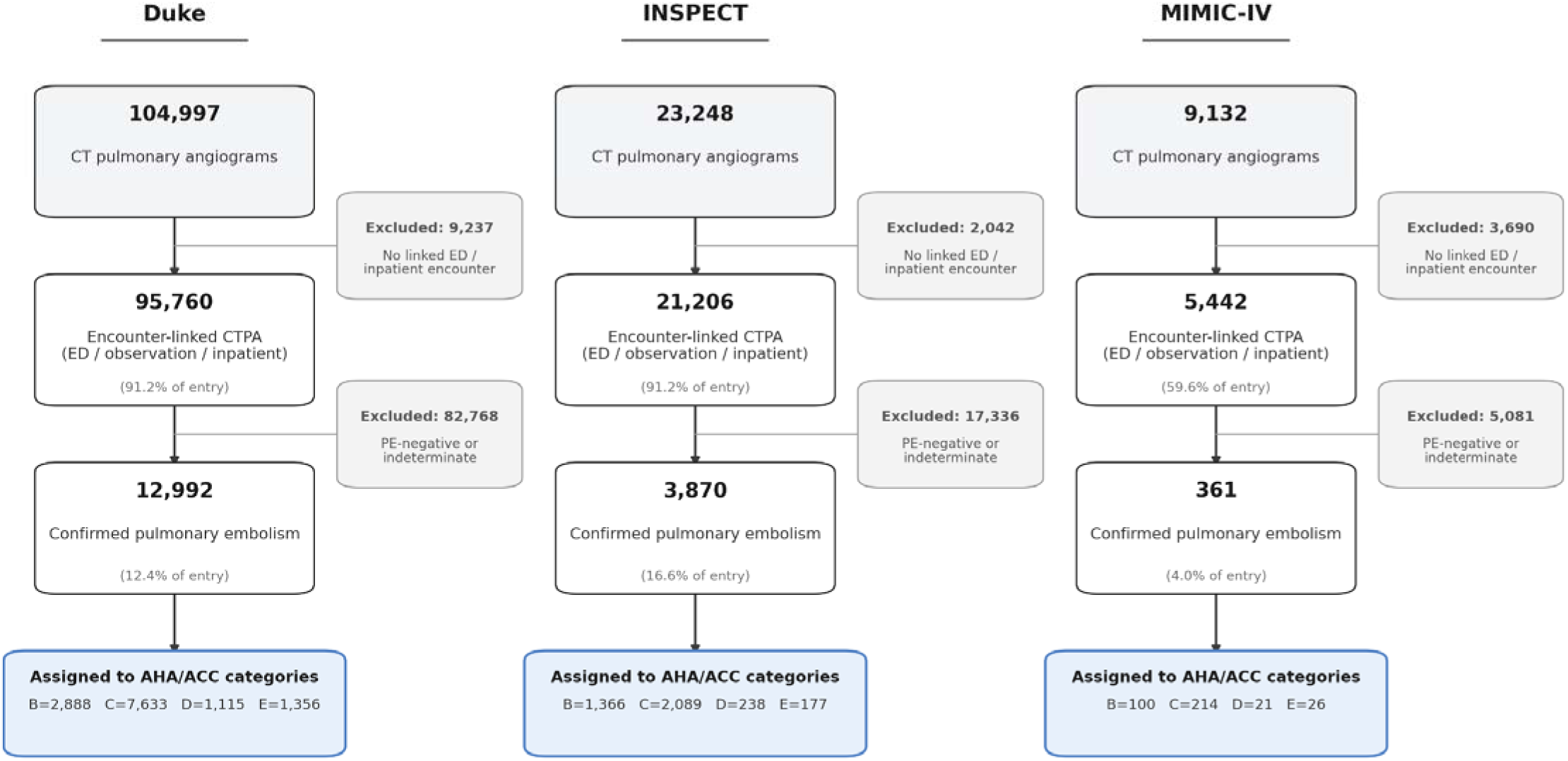
Derivation of the study cohorts. Flow of patients from 95,760 consecutive inpatient CTPA studies at Duke, through diagnostic exclusions, to 12,992 patients with confirmed PE assigned to AHA/ACC clinical categories, with the INSPECT (n=3,870) and MIMIC-IV (n=361) cohorts shown as parallel derivations. TPA indicates computed tomographic pulmonary angiography; PE, pulmonary embolism. Datasets: Duke (primary), INSPECT, MIMIC-IV.

**Table 1.**
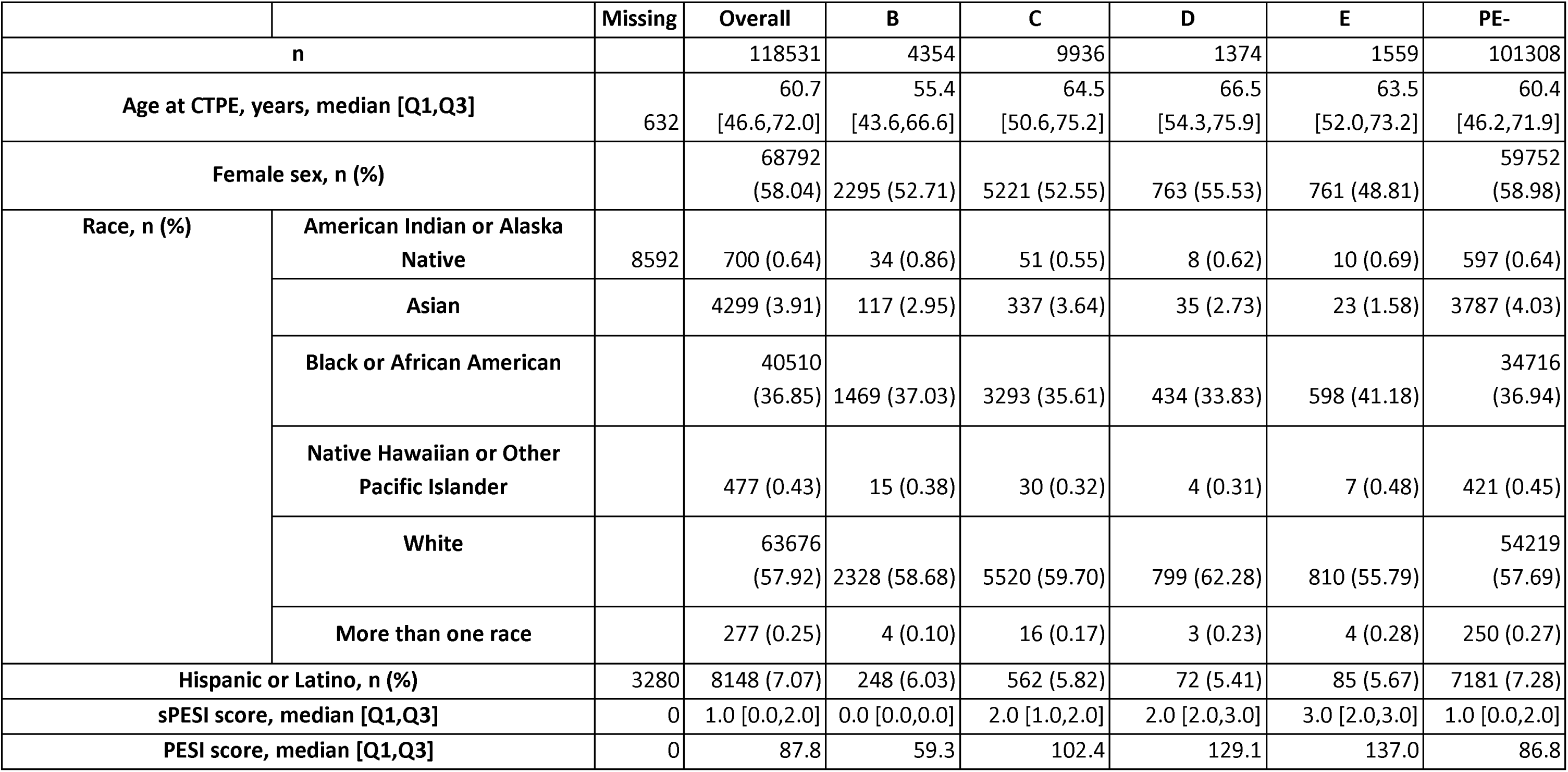
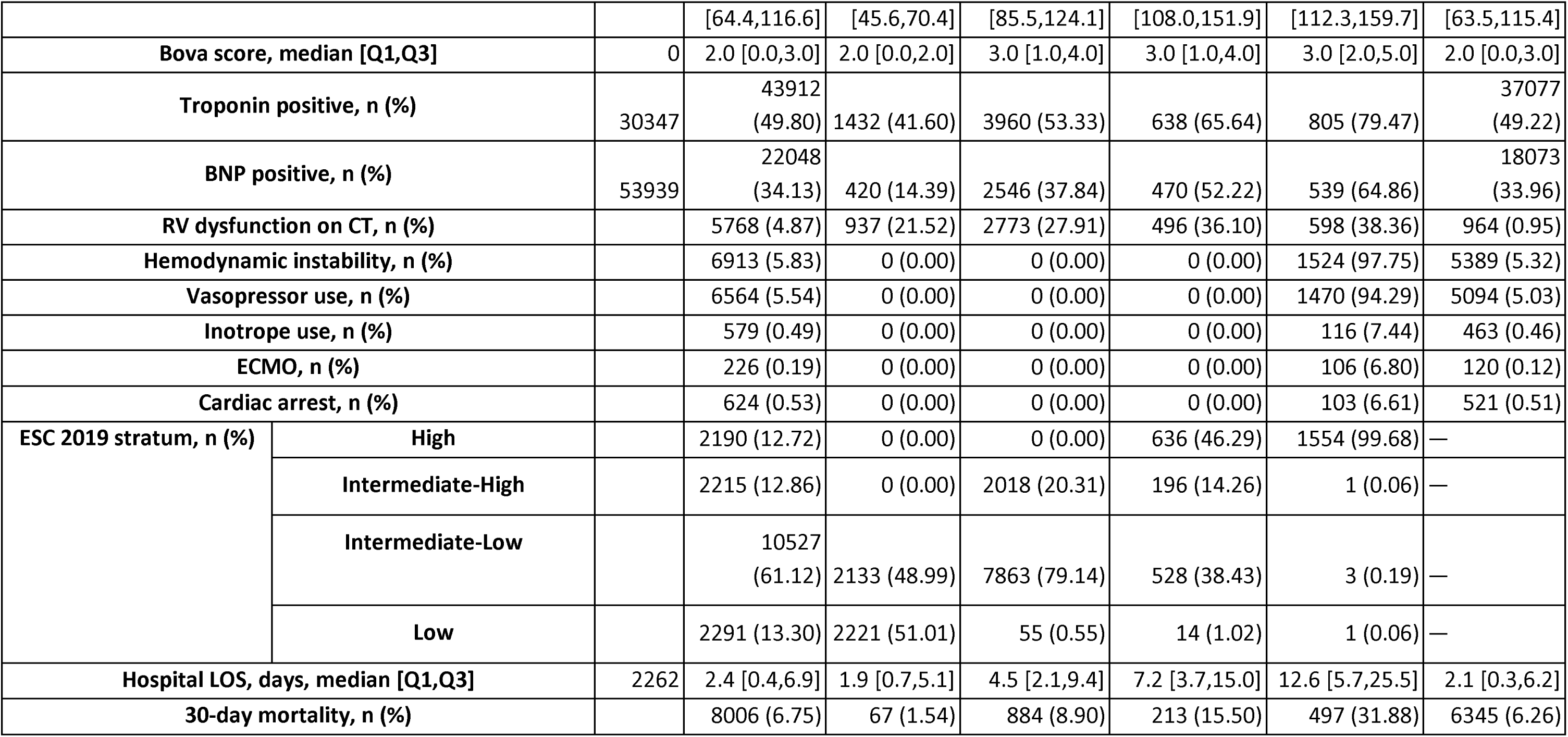
Baseline characteristics of patients with and without pulmonary embolism, by AHA/ACC clinical category, in three health systems. Characteristics are reported for the Duke (n=12,992), INSPECT (n=3,870), and MIMIC-IV (n=361) cohorts of the and are stratified by 2026 AHA/ACC clinical category (B through E). This also reflects a pooled PE negative cohort (PE-negative) to contrast against the PE+, including continuous PE-. Note that PE- from INSPECT and MIMIC are not known to be consecutive. Categorical variables are reported as n (%) and continuous variables as median (IQR). ACC indicates American College of Cardiology; AHA, American Heart Association; IQR, interquartile range; PE, pulmonary embolism; PESI, PE severity index; BNP, b-type natriuretic peptide; RV, right ventricle; LOS, length of stay.

The Duke cohort was derived from 95,760 consecutive inpatient CTPA studies, of which 12,992 (13.6%) were positive for PE; the remaining 82,768 PE-negative studies constituted a consecutive imaging denominator not available in the other two external cohorts. This permitted analysis of positivity rates by site, which yielded a range of 7.7% to 11.0% at DUHS community hospitals and 15.6% at the quaternary academic center. (**Table S23**)

Markers of severity increased monotonically independently across the B-E categories (pooled **Table 1**; per-site **Table S1a-S1c)**. In the pooled cohort, the median sPESI rose from 0 to 3 and the median PESI score from 59 to 137. RV dysfunction on CT increased from 21.5% (B) through 38.3% (E), troponin positivity from 41.6% (B) to 79.5% (E), and B-type natriuretic positivity from 14.4% (B) to 64.9% (E). **(Table S1)** Category E was defined by hemodynamic instability (97.8%) as demonstrated by either vasopressors (94.3%), extracorporeal membrane oxygenation (6.8%), or cardiac arrest (6.6%), which were not mutually exclusive.

RV dysfunction was identified by CT in 31.3% (4,064/12,992), echocardiography in 30.0% (3,900/12,992), and in both CT and echocardiography in only 16% (2,080/12,992); 45.3% (5,884/12,992) had RV dysfunction on at least one modality. These patients were treated as C1, though RV dysfunction reclassifying to C2 cannot be excluded.

Thirty-day mortality among the imaged but PE-negative pooled cohort population was 6.3%, exceeding that of category B (1.5%), demonstrating that the lowest-acuity confirmed PE carries lower short-term mortality than the comorbidity burden of the population in whom PE is suspected but not confirmed.

### Outcomes by Clinical Category

Thirty-day mortality rose across categories in the pooled cohort: 1.5% (B), 8.9% (C), 15.5% (D), and 31.9% (E). To test whether this gradient was true across all cohorts, we examined each cohort separately. The same ordering held at Duke: 1.6% (B), 9.5% (C), 14.8% (D), and 32.7% (E), INSPECT (B: 1.5%, C: 6.7%, D: 19.3%, E: 26.0%), and in MIMIC-IV (B: 0.0%, C: 8.4%, D: 9.5%, E: 30.8%), reproducing across cohorts with a range of baseline mortality (**Figure 2, Table S2**). The ordering was preserved for in-hospital and one-year mortality (**Table S2**).

**Figure 2.**
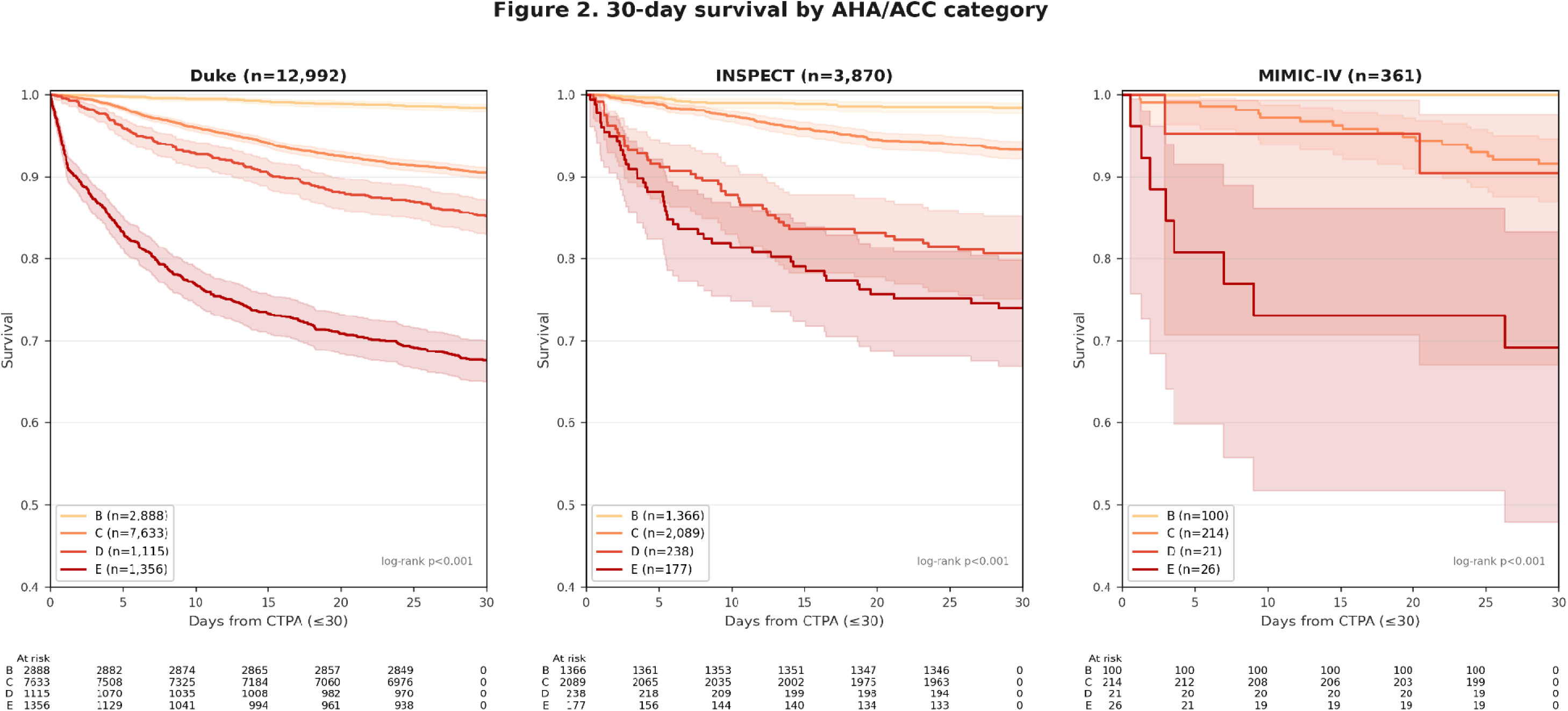
Thirty-day survival by AHA/ACC clinical category in three health systems. Kaplan-Meier survival curves for AHA/ACC categories B through E are shown in separate panels for the Duke, INSPECT, and MIMIC-IV cohorts. Numbers at risk and 30-day mortality are annotated beneath each panel. Estimates in MIMIC-IV are limited by sample size. Datasets: Duke, INSPECT, MIMIC-IV.

#### Subcategory Resolution

Across the two cohorts (Duke, INSPECT) with sufficient patients to evaluate subcategory differences with confidence intervals, separation of mortality emerged reliably in subcategories D2-E2. The intermediate band C1-D1 subcategories had differing utility across cohorts. At Duke, thirty-day mortality was similar across the C1-D1 categories, varying by less than 3.0 percentage points (8.1% in C3 to 10.9% in D1), and no monotonic trend was present across the ordered subcategories. (Cochran-Armitage z=-0.204, p=0.8382; Cox hazard ratio per step 0.99, p=0.88, **Table S6**) Subcategories C1-D1 added no discrimination over treating C1-D1 as a single group (Harrell C-index 0.696 vs 0.697; difference -0.001) (**Table S6**). In INSPECT, a modest ordered gradient was present between C1-D1 (Cochran-Armitage z=3.121, p=0.0018; Cox hazard ratio per step 1.28, 95% CI 1.10-1.49), and the full structure added modest discrimination (C-index difference +0.03), driven by separation of D1 instead of an ordered mortality increase between C1-C3. (**Figure 3, Table S6**) Of note, INSPECT did not contain echocardiography data and so captured the presence of RV strain by CT alone, missing patients with TTE-only RV strain.

**Figure 3.** Thirty-day mortality and advanced-therapy use across AHA/ACC subcategories, Duke cohort. A two-panel figure sharing a common subcategory axis. Panel A shows 30-day mortality for subcategories B1 through E2 as points with 95% Wilson confidence intervals. Panel B shows the proportion of patients receiving each therapy (anticoagulation alone, systemic thrombolysis, catheter-directed thrombolysis, mechanical thrombectomy, and extracorporeal membrane oxygenation), aligned beneath Panel A. The figure is restricted to the Duke cohort because advanced-therapy data are available only at Duke, and pairing mortality with treatment within a single cohort is required for the escalation comparison. Reproduction of the subcategory mortality pattern in INSPECT is shown in Figure S1 and stated in the Results text. Datasets: Duke.

Thirty-day mortality at Duke was 0% (B1, 0/136), 1.7% (B2, 46/2,752), 9.2% (C1, 219/2,380), 10.8% (C2, 326/3,015), 8.1% (C3, 181/2,238), 10.9% (D1, 85/783), 24.1% (D2, 80/332), 31.0% (E1, 375/1,208), and 45.9% (E2, 68/148) (**Figure 3**, **Table S2**). Pairwise differences within C1-D1 were small, except for C3, which was lower than both C2 (difference +2.73 percentage points [pp], 95% CI 1.12-4.29) and D1 (-2.77pp, 95% CI - 5.37- 0.44; **Table S5**). To test whether the four subcategories carried information beyond a single pooled band, we compared nested Cox models; likelihood-ratio test favored the four-level C1-D1 model over a single pooled-band indicator (p=0.006), reflecting the lower C3 estimate rather than an ordered gradient, and the C3 hazard violated proportionality over the 30-day window (Schoenfeld < 0.001). (**Table S6**) C3 also carried the lowest mortality at 1 year (24.4%) between C1-D1 subcategories **(Table S2)**. The flat-band result was insensitive to the handling of C-indeterminate (no RV dysfunction on CT with no TTE or biomarkers measured) patients. (**Table S9**)

### C-Indeterminate

A subset of category C patients could not be subclassified. At Duke, 967 of 7,633 category C patients (12.7%) had neither echocardiography nor cardiac biomarkers within the ascertainment window and were designated “*C-indeterminate”*; the corresponding proportion in INSPECT was 643 of 2,089 (30.8%). Thirty-day mortality in this group was 10.4% (95% CI 8.7–12.5) at Duke and 7.3% (95% CI 5.5–9.6) in INSPECT, equal to or exceeding that of the subclassified C1–C3 patients rather than falling below them. Excluding these patients, treating them as a separate stratum, or reassigning them to C1 left the intermediate-band mortality pattern unchanged **(Table S9**), indicating that the flat-band result does not depend on their handling. The absence of subclassifying data does not identify a lower-risk subgroup.

### Treatment

Treatment data was consistently available only for the Duke data. At Duke, advanced therapy was concentrated in the high-acuity subcategories (**Figure 3, Table S4)**. Anticoagulation alone predominated across all categories, and was used in >90% of cases through C2. Aspiration/mechanical thrombectomy (n=468, 3.6%), non-ultrasound catheter directed thrombolytics (n=137,1.1%) were rare overall but increased across subcategories. Systemic thrombolysis was given in 311 (2.4%), concentrated in categories E1-E2. We were unable to differentiate between surgical embolectomies and endarterectomies, and as such do not report these numbers.

#### Comparison with the 2019 ESC strata

The frameworks were partially concordant at the extremes and divergent in the intermediate-low and intermediate-high categories. (**Figure 4, Tables S10-S13**) At Duke, 94.7% of ESC low-risk patients mapped to category B (1,142/1,206), and the majority of ESC high-risk patients mapped to Category E (74.9%, 1,352/1,804) and the rest mapped to Category D (25.1%, 452/1,804). Almost 25% of ESC intermediate-low were translated to category B (21.9%, 1,746/7,956). The ESC intermediate-low and intermediate-high strata were redistributed: 72.3% (5,752/7,956) of intermediate-low and 90.3% (1,830/2,026) of intermediate-high patients remained in category C, while 5.7% (456/7,956) and 9.6% (195/2,026) respectively were reclassified to category D. Patients reclassified to D carried modestly higher 30-day mortality that did not reach statistical significance (11.6% vs 9.8% among intermediate-low patients, risk difference +1.8pp, 95% CI -1-5.1 and intermediate-high: 9.7% vs 8.6%, +1.2pp, 95% CI -2.5-6.3). (**Table S11**)

**Figure 4.**
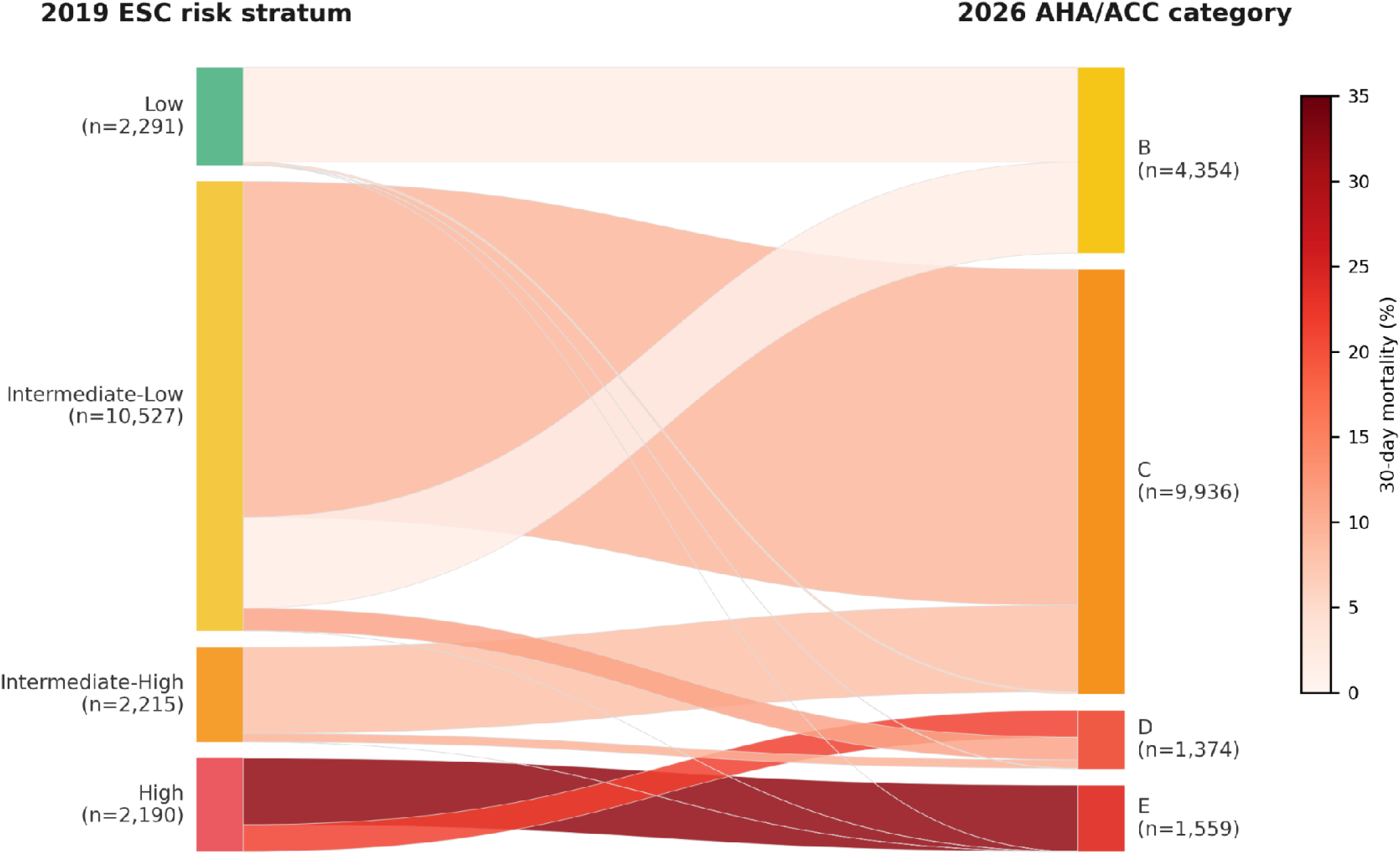
Reclassification of patients from 2019 ESC risk strata to 2026 AHA/ACC clinical categories. Alluvial diagram of patient flow from ESC risk strata (low, intermediate-low, intermediate-high, high) to AHA/ACC categories (B through E) across all cohorts, with flows sized by the number of patients and shaded by 30-day mortality. ESC indicates European Society of Cardiology. Datasets: Duke, INSPECT, MIMICIV.

## Discussion

In 17,223 patients with confirmed PE across three academic health systems, the 2026 AHA/ACC framework produced a reliable, reproducible mortality gradient at the category level but added little prognostic resolution across the broad intermediate band (C1-D1). To our knowledge, this is the first multi-cohort validation of these categories against clinical outcomes, a gap the guideline’s writing committee identified as a priority, particularly for artificial intelligence-based approaches.^9^ Ordering was preserved in all three cohorts, indicating the gradient reflects the categories rather than differences in case mix. The added subcategory granularity refined risk primarily at higher acuities D2-E2, where treatment escalation decisions are often made; across the C1-D1 band, subcategories did not separate.

### An ordinal mortality gradient reproducible across cohorts

Thirty-day mortality increased monotonically from category B through E, and in-hospital and one-year mortality demonstrated similar ordering, with progressive and sustained separation through one year. The increase per category grew more steeply at higher acuity categories, with the transition to Category E as the largest single increment. This mortality gradient order was preserved in MIMIC-IV and INSPECT as well.Prior national PE registries stratified by ESC criteria have likewise shown similar survival across the lower-risk strata with a steep decline at the high-risk extreme.^50^ Reproducible ordering across cohorts supports the use of the categories for relative risk stratification across populations, while indicating that absolute risk estimates remain population-dependent. Unlike registry-based cohorts, the DUHS cohort was assembled from all consecutive CTPA studies and therefore reflects a real-world, unselected population evaluated for suspected PE — a feature shared to a lesser extent by MIMIC-IV and INSPECT — rather than the selected populations underlying most prior evidence. As the categories were set by expert consensus rather than derived from outcomes, their reproducible association with 30-day mortality in an unselected population and two external cohorts is direct evidence that the categories track risk.

### Added granularity refines risk principally at the high-acuity end

The subcategories added prognostic information primarily at the high-acuity end. Separation emerged only within D2, shock (E1), and refractory cardiogenic shock or cardiac arrest (E2), categories that the guideline recommends advanced intervention like mechanical thrombectomy.

Two observations within category C warrant further emphasis. First, the C subcategories did not order monotonically by mortality. In the Duke cohort, thirty-day mortality was 9.2% in C1, 10.8% in C2, and 8.1% in C3. The observation that C3, the subcategory defined by the most abnormal physiology, carried the lowest short-term mortality is counterintuitive if these markers are interpreted as a simple severity axis. A plausible explanation is treatment-related, as C3 patients received advanced therapy more often than C1 or C2 (catheter-directed thrombolysis in 2.8% and aspiration thrombectomy in 6.6% in C3), whereas nearly all C1 patients received anticoagulation alone. This pattern was seen in HI-PEITHO and STORM-PE, with advanced therapies reducing decompensation and patients in the control arm also treated as they deteriorated, with markers functioning less like independent prognosticators than as triggers for more advanced therapies.^51,52^ The markers may therefore be performing as intended, prompting escalation that attenuates the risk they identify. Second, the overlap among the C subtiers, and of D1 with C, indicates that the discriminative information added by the subcategories is realized at the D2 and E boundaries, where treatment decisions are most consequential, rather than within the intermediate subdivisions. The pattern was reproduced in INSPECT (where RV was defined only by CT, as echocardiography was not available) and, in the smaller, fully phenotyped MIMIC-IV cohort, although both external cohorts were underpowered for stratum level inference.

### Category C patients who cannot be subclassified are not a low-risk group

A substantial proportion of category C patients (967/7,633 or 12.7% of the category C patients), could not be assigned to a subcategory because they lacked both echocardiography and biomarker testing — data the AHA/ACC 2026 framework requires for C subcategorization because they were judged important to risk assessment.(**Table S9**) Their 30-day mortality (9.2%) equalled or exceeded that of the fully classified C1–C3 patients, contradicting the assumption that missing tests reflect a lower-risk presentation not warranting workup. The Duke cohort captures every order placed, so this pattern reflects a gap in clinical practice rather than a gap in data capture: the echocardiograms and biomarkers were not performed, not merely unrecorded. This finding does not exclude the possibility that point-of-care ultrasound was used to guide management outside the structured record. Any framework that subclassifies on tests that are not uniformly obtained will leave a portion of patients indeterminate at the bedside, and their outcomes indicate that they should not be presumed to be at low risk. This gap is in principle modifiable — a target for future quality-improvement or education efforts rather than a fixed limitation of the framework.

### Reclassification and treatment implications

When mapped against the 2019 ESC scheme, the AHA/ACC framework redistributed patients almost entirely within the intermediate strata while leaving the extremes concordant, a comparison the AHA 2026 guidelines invite.^30,31^ ESC low-risk patients became category B in 94.7% of cases (1142/1,206), and the majority of ESC high-risk patients became category E (74.9%, 1,352/1,804); category E was in turn drawn almost exclusively from ESC high risk (99.7%, 1,352/1,356). In contrast, the ESC intermediate-low and intermediate-high groups dispersed, with the framework separating a hemodynamically defined subgroup to category D from the larger group in category C. The principal reorganizing effect of the new system is therefore to re-partition the heterogeneous ESC intermediate population.

Whether this re-partitioning alters management is a separate question. Treatment intensity tracked category as expected. Anticoagulation alone declined from 98.1% in category B to 82.0% in category E, and systemic thrombolysis was most frequent in category E (9.9%). That being said, systemic thrombolysis was far less frequent in this cohort than in previous literature. ^53–56^ Advanced therapies nonetheless remained uncommon even at the highest tiers, and most patients in every category received anticoagulation alone.

### Limitations

The AHA/ACC categories are expert-consensus constructs not derived from or validated against outcomes, applied here retrospectively to data not collected for this purpose; this analysis evaluates their prognostic behavior without establishing that the boundaries are optimally placed.^9^ The flat intermediate band (C1-D1) cannot distinguish limited prognostic resolution from a gradient attenuated by treatment, an ambiguity inherent to the rescue-on-deterioration design. Cox models entered the classification as the sole term, so hazard ratios reflect discrimination rather than a treatment-independent effect, and escalation to advanced therapy, more frequent in the higher subcategories, was not adjusted for. Proportional hazards were violated for C3, making its hazard ratio a time-averaged summary. Within-band comparisons were exploratory and uncorrected for multiplicity, with equivalence margins of 3.0 and 2.0 percentage points prespecified rather than empirically derived. Category assignment was complete-case, treating an unobtained test as a criterion not met, which biases toward lower acuity.

Assignment also depended in part on LLM extraction, and misclassification of imaging findings would propagate into subcategories.^57^ Category E warrants particular caution, because instability ascertained from vasopressor administration and cardiac-arrest flowsheets cannot be attributed to PE with certainty. E1 likely includes shock from competing causes such as sepsis, whereas E2 captures the most extreme presentations, which may explain the higher mortality in E2 (45.9%) than E1 (31.0%). Mortality in the highest-acuity tier therefore overstates PE-specific lethality, which may also explain why so many category E patients received no intervention beyond anticoagulation. The C-indeterminate group, the relationship between categories C and D, and the R modifier each merit dedicated investigation.

### Conclusions

In a denominator-level, multi-system cohort, the 2026 AHA/ACC framework stratified short-term mortality in a reproducible, ordinal fashion at the category level. Subcategory granularity refined risk principally at the high-acuity end (D2 and E), but the C and D1 subtiers overlapped and did not order monotonically.The reorganizing effect of the framework relative to the 2019 ESC scheme falls almost entirely on the intermediate strata. These findings support the discriminative validity of the new categories and identify specific opportunities for refinement, including reconciliation of the orthogonal C and D axes, recognition of physiologically abnormal category B patients, and management of the unclassifiable C-indeterminate group.

## Data Availability

All data produced in the present study are contained in the supplemental file and further is available upon reasonable request to the authors.

## Conflicts of interest

AIW holds equity and management positions in Ataia Medical, AI Wong Consulting, LLC, and Synappsed, LLC, and serves as a consultant for Merck Sharp & Dohme, LLC. AIW is supported by the National Heart, Lung, and Blood Institute (NHLBI; 1R01HL177003, 5R01HL176569), the National Institute of General Medical Sciences (NIGMS; 1R33GM146142), and the National Institute on Minority Health and Health Disparities (NIMHD; U54MD012530, REACH Equity).

JGM serves as a consultant for Jupiter Endovascular, Penumbra Inc, Inari Medical Inc and Becton Dickinson and Co.

SR has received consulting fees from Johnson & Johnson, Gossamer Bio, Inhibikase Therapeutics, Inc., Insmed, Liquidia Technologies, Inc., Merck & Co, Inc., Pahr Therapeutics, Pulmovant, and United Therapeutics Corp., has a patent licensed by Polarean,, Inc., and has received research grants from the American Heart Association, National Institutes of Health and the Heart, Lung and Blood Institute, Johnson & Johnson, MSD, and United Therapeutics Corp.

WSJ: Research grants to DCRI that support salary: Bayer, Boehringer Ingelheim, Merck, Novartis, National Institutes of Health, Patient-Centered Outcomes Research Institute, Regeneron. Consulting/Advisory roles: American College of Physicians, Amplitude Vascular Solutions, General Electric Healthcare, Merck, U.S. Department of Justice

NLBF receives research support from the National Institutes of Health, the American Diabetes Association, Novartis, and Amgen, none of which supported the present study.

VT serves as Sr. Director, Medical Affairs, Stryker PV.

TO: Research Support: Stago, Werfen, Takeda, Octapharma, Anthos, CDC, and NIH. Consultant positions: Sanofi, Genentech. Royalties: Up To Date

All other authors report no conflicts of interest.

## Acknowledgements

The authors thank Willard Applefeld, MD, for his contributions to the development of the Duke Pulmonary Embolism Response Team pathway.

