## Supplemental Tables and FIgures for "Real-World Performance of the 2026 AHA/ACC Pulmonary Embolism Framework in a Multi-System CTPA Cohort"

### Supplemental text

Despite the clinical importance of PE, our understanding of optimal management remains limited by challenges in data capture and integration.^57^ Much of the critical information (*e.g.*, exact clot location, presence of right heart strain) is documented in semi-structured free-text radiology reports or cardiology notes, making them heterogeneous and not readily analyzable.^57^ Registries, such as the Pulmonary Embolism Response Team (PERT) Consortium registry, have been developed to standardize and study PE care. Still, they typically require resource-intensive manual abstraction of data from charts and reports, which limits broad adoption.^17,57^ Emerging artificial intelligence (AI) technologies, particularly large language models (LLMs), offer promising solutions by converting unstructured clinical narratives into structured, analyzable data formats.^19^ Thus, comprehensive management of PE relies on integrating multi-modal data: imaging findings (PE location, clot burden, right heart strain), physiological data (*e.g.,* vital signs, EKG changes), laboratory results, and clinical context.

#### Dataset descriptor

Radiology text reports for CTPE studies, along with the corresponding original CTPE images, were retrieved and stored in ASCII text (from Epic Clarity) and DICOM (from the vendor neutral archive). format. The free-text radiology reports were processed using an LLM-based information extraction pipeline, in accordance with structured data concepts from the PERT Consortium,^57^ to identify structured PE-related concepts as per Alwakeel et al 2026.^18^ The LLM was prompted to extract key findings from each report, including (a) whether an acute PE was present or absent, (b) the anatomical location of any detected emboli (*e.g.*, main pulmonary artery, lobar, segmental, subsegmental branches), (c) indicators of right heart strain on the CT (such as right ventricular enlargement or interventricular septal bowing), and (d) the presence of significant ancillary findings or image quality issues (*e.g.*, motion artifacts).

We obtained both the original images (in DICOM format) and the corresponding text reports from transthoracic echocardiograms, as well as EKG waveforms in GE MUSE (GE Healthcare, Waukesha, WI) XML format, converted to WFDB, and their interpretations (from both computer-generated and physician-provided readings). Echocardiography and EKG data are available in this dataset as both imaging files and narrative reports, allowing for the future development and validation of imaging-based machine learning models. The overall dataset was assembled by securely linking the above data sources using unique study identifiers and encounter dates.

### Supplemental Tables

#### Supplemental Table S1. Complete baseline characteristics by AHA/ACC clinical category by cohort.

Demographic, comorbidity, vital-sign, laboratory, and imaging variables are reported by cohort stratified by AHA/ACC clinical category (B through E). Categorical variables are reported as n (%) and continuous variables as median (IQR). ACC indicates American College of Cardiology; AHA, American Heart Association; BNP, B-type natriuretic peptide; CT, computed tomography; CTPA, computed tomography pulmonary angiography; ECMO, extracorporeal membrane oxygenation; ESC, European Society of Cardiology; INSPECT, Integrating Numerous Sources for Prognostic Evaluation of Clinical Timelines; IQR, interquartile range; LOS, length of stay; MAP, mean arterial pressure; MIMIC-IV, Medical Information Mart for Intensive Care IV; PE, pulmonary embolism; PESI, Pulmonary Embolism Severity Index; Q1 and Q3, first and third quartiles; RV, right ventricular; SBP, systolic blood pressure; SpO2, peripheral oxygen saturation; sPESI, simplified Pulmonary Embolism Severity Index; TTE, transthoracic echocardiography.

##### Table S1a. Duke Cohort

| **Characteristic** | **Missing** | **Overall** | **B** | **C** | **D** | **E** | **PE-** |
| --- | --- | --- | --- | --- | --- | --- | --- |
| **n** |  | 95760 | 2888 | 7633 | 1115 | 1356 | 82768 |
| **Age, years, median [Q1,Q3]** | 0 | 60.5 [46.3,71.9] | 54.9 [43.1,66.5] | 64.6 [50.4,75.2] | 67.4 [55.2,76.4] | 63.6 [52.2,73.3] | 60.1 [45.8,71.6] |
| **Female sex, n (%)** |  | 55893 (58.37) | 1554 (53.81) | 3998 (52.38) | 629 (56.41) | 658 (48.53) | 49054 (59.27) |
| **Race, n (%) — American Indian or Alaska Native** | 4129 | 631 (0.69) | 28 (1.01) | 46 (0.62) | 7 (0.65) | 10 (0.77) | 540 (0.68) |
| **Race, n (%) — Asian** |  | 1374 (1.50) | 25 (0.90) | 78 (1.06) | 14 (1.30) | 8 (0.62) | 1249 (1.58) |
| **Race, n (%) — Black or African American** |  | 38458 (41.97) | 1354 (48.78) | 3119 (42.35) | 420 (39.07) | 577 (44.52) | 32988 (41.69) |
| **Race, n (%) — Native Hawaiian or Other Pacific Islander** |  | 112 (0.12) | 1 (0.04) | 5 (0.07) | 0 (0.00) | 0 (0.00) | 106 (0.13) |
| **Race, n (%) — White** |  | 50780 (55.42) | 1364 (49.14) | 4100 (55.68) | 631 (58.70) | 697 (53.78) | 43988 (55.60) |
| **Race, n (%) — More than one race** |  | 276 (0.30) | 4 (0.14) | 16 (0.22) | 3 (0.28) | 4 (0.31) | 249 (0.31) |
| **Hispanic or Latino, n (%)** | 2527 | 4784 (5.13) | 123 (4.41) | 258 (3.48) | 36 (3.33) | 52 (3.97) | 4315 (5.35) |
| **sPESI score, median [Q1,Q3]** | 0 | 1.0 [0.0,2.0] | 0.0 [0.0,0.0] | 1.0 [1.0,2.0] | 2.0 [2.0,3.0] | 3.0 [2.0,3.0] | 1.0 [0.0,2.0] |
| **PESI score, median [Q1,Q3]** | 0 | 87.9 [64.8,116.7] | 61.5 [48.0,72.1] | 103.2 [86.7,125.0] | 127.9 [108.0,151.2] | 136.4 [110.9,159.0] | 86.4 [63.5,115.2] |
| **Bova score, median [Q1,Q3]** | 0 | 2.0 [0.0,3.0] | 2.0 [0.0,2.0] | 3.0 [1.0,4.0] | 3.0 [2.0,5.0] | 3.0 [2.0,5.0] | 2.0 [0.0,3.0] |
| **Charlson index, median [Q1,Q3]** | 0 | 0.0 [0.0,2.0] | 0.0 [0.0,0.0] | 1.0 [0.0,2.0] | 1.0 [0.0,2.0] | 1.0 [0.0,2.0] | 0.0 [0.0,2.0] |
| **Cancer, n (%)** |  | 14675 (15.32) | 0 (0.00) | 2006 (26.28) | 246 (22.06) | 237 (17.48) | 12186 (14.72) |
| **Cardiopulmonary disease, n (%)** |  | 15731 (16.43) | 0 (0.00) | 1262 (16.53) | 196 (17.58) | 275 (20.28) | 13998 (16.91) |
| **Heart failure, n (%)** |  | 6913 (7.22) | 0 (0.00) | 591 (7.74) | 116 (10.40) | 209 (15.41) | 5997 (7.25) |
| **Chronic lung disease, n (%)** |  | 10044 (10.49) | 0 (0.00) | 760 (9.96) | 93 (8.34) | 83 (6.12) | 9108 (11.00) |
| **Heart rate (max), median [Q1,Q3]** | 1858 | 102.0 [88.0,119.0] | 92.0 [83.0,101.0] | 111.0 [96.0,124.0] | 113.0 [98.0,130.0] | 126.0 [109.0,143.0] | 102.0 [87.0,118.0] |
| **SBP (min), median [Q1,Q3]** | 2183 | 110.0 [98.0,124.0] | 116.0 [108.0,126.0] | 106.0 [97.0,118.0] | 85.0 [77.0,94.0] | 81.0 [69.0,94.5] | 111.0 [99.0,125.0] |
| **MAP (min), median [Q1,Q3]** | 16186 | 79.0 [68.0,90.0] | 85.0 [77.0,94.0] | 78.0 [70.0,87.0] | 55.0 [48.0,58.0] | 52.0 [40.0,61.0] | 79.0 [69.0,91.0] |
| **Resp rate (max), median [Q1,Q3]** | 1970 | 22.0 [20.0,28.0] | 21.0 [19.0,24.0] | 24.0 [20.0,29.0] | 28.0 [23.0,32.0] | 33.0 [28.0,40.0] | 22.0 [20.0,28.0] |
| **SpO2 (min), median [Q1,Q3]** | 1888 | 94.0 [91.0,97.0] | 95.0 [93.0,97.0] | 93.0 [90.0,95.0] | 91.0 [88.0,94.0] | 89.0 [82.0,92.0] | 94.0 [91.0,97.0] |
| **Temp (min), median [Q1,Q3]** | 2305 | 36.5 [36.3,36.7] | 36.5 [36.4,36.7] | 36.4 [36.3,36.6] | 36.4 [36.3,36.6] | 36.3 [35.7,36.5] | 36.5 [36.3,36.7] |
| **Troponin positive, n (%)** | 30347 | 40541 (61.98) | 1180 (59.72) | 3403 (66.43) | 552 (77.42) | 730 (90.12) | 34676 (61.06) |
| **BNP positive, n (%)** | 53939 | 19631 (46.94) | 398 (27.39) | 2199 (49.69) | 414 (64.59) | 496 (78.98) | 16124 (46.50) |
| **Lactate >2 mmol/L, n (%)** |  | 10369 (10.83) | 84 (2.91) | 915 (11.99) | 279 (25.02) | 753 (55.53) | 8338 (10.07) |
| **Creatinine rise, n (%)** |  | 3222 (3.36) | 49 (1.70) | 213 (2.79) | 75 (6.73) | 259 (19.10) | 2626 (3.17) |
| **RV dysfunction on CT, n (%)** |  | 4548 (4.75) | 764 (26.45) | 2332 (30.55) | 435 (39.01) | 533 (39.31) | 484 (0.58) |
| **RV dysfunction on TTE, n (%)** |  | 15255 (15.93) | 537 (18.59) | 2295 (30.07) | 415 (37.22) | 653 (48.16) | 11355 (13.72) |
| **Subsegmental-only clot, n (%)** |  | 591 (0.62) | 136 (4.71) | 338 (4.43) | 57 (5.11) | 60 (4.42) | 0 (0.00) |
| **Hemodynamic instability, n (%)** |  | 5781 (6.04) | 0 (0.00) | 0 (0.00) | 0 (0.00) | 1343 (99.04) | 4438 (5.36) |
| **Vasopressor, n (%)** |  | 5539 (5.78) | 0 (0.00) | 0 (0.00) | 0 (0.00) | 1303 (96.09) | 4236 (5.12) |
| **Inotrope, n (%)** |  | 419 (0.44) | 0 (0.00) | 0 (0.00) | 0 (0.00) | 89 (6.56) | 330 (0.40) |
| **ECMO, n (%)** |  | 185 (0.19) | 0 (0.00) | 0 (0.00) | 0 (0.00) | 83 (6.12) | 102 (0.12) |
| **Cardiac arrest, n (%)** |  | 369 (0.39) | 0 (0.00) | 0 (0.00) | 0 (0.00) | 68 (5.01) | 301 (0.36) |
| **ESC 2019 stratum, n (%) — Low** |  | 1206 (9.28) | 1142 (39.54) | 51 (0.67) | 12 (1.08) | 1 (0.07) | — |
| **ESC 2019 stratum, n (%) — Intermediate-Low** |  | 7956 (61.24) | 1746 (60.46) | 5752 (75.36) | 456 (40.90) | 2 (0.15) | — |
| **ESC 2019 stratum, n (%) — Intermediate-High** |  | 2026 (15.59) | 0 (0.00) | 1830 (23.97) | 195 (17.49) | 1 (0.07) | — |
| **ESC 2019 stratum, n (%) — High** |  | 1804 (13.89) | 0 (0.00) | 0 (0.00) | 452 (40.54) | 1352 (99.71) | — |
| **Hospital LOS, days, median [Q1,Q3]** | 0 | 2.3 [0.4,6.6] | 1.9 [0.7,5.0] | 4.3 [2.1,9.1] | 6.9 [3.5,13.9] | 12.4 [5.7,25.5] | 2.0 [0.3,6.0] |
| **In-hospital mortality, n (%)** |  | 3311 (3.46) | 19 (0.66) | 251 (3.29) | 77 (6.91) | 386 (28.47) | 2578 (3.11) |
| **30-day mortality, n (%)** |  | 6608 (6.90) | 46 (1.59) | 726 (9.51) | 165 (14.80) | 443 (32.67) | 5228 (6.32) |
| **1-year mortality, n (%)** |  | 17993 (18.79) | 200 (6.93) | 2010 (26.33) | 380 (34.08) | 633 (46.68) | 14770 (17.85) |

##### Table S1b. INSPECT Cohort

| **Characteristic** | **Missing** | **Overall** | **B** | **C** | **D** | **E** | **PE-** |
| --- | --- | --- | --- | --- | --- | --- | --- |
| **n** |  | 17869 | 1366 | 2089 | 238 | 177 | 13999 |
| **Age, years, median [Q1,Q3]** | 632 | 60.7 [46.9,72.0] | 56.9 [45.1,66.8] | 64.3 [51.5,75.0] | 63.5 [51.8,72.8] | 62.3 [48.0,71.8] | 60.5 [46.4,71.9] |
| **Female sex, n (%)** |  | 10151 (56.81) | 684 (50.07) | 1091 (52.23) | 119 (50.00) | 89 (50.28) | 8168 (58.35) |
| **Race, n (%) — American Indian or Alaska Native** | 3821 | 64 (0.46) | 6 (0.55) | 5 (0.29) | 1 (0.53) | 0 (0.00) | 52 (0.48) |
| **Race, n (%) — Asian** |  | 2735 (19.47) | 89 (8.12) | 250 (14.74) | 21 (11.11) | 15 (11.03) | 2360 (21.59) |
| **Race, n (%) — Black or African American** |  | 1165 (8.29) | 92 (8.39) | 136 (8.02) | 11 (5.82) | 18 (13.24) | 908 (8.31) |
| **Race, n (%) — Native Hawaiian or Other Pacific Islander** |  | 360 (2.56) | 14 (1.28) | 25 (1.47) | 4 (2.12) | 7 (5.15) | 310 (2.84) |
| **Race, n (%) — White** |  | 9724 (69.22) | 895 (81.66) | 1280 (75.47) | 152 (80.42) | 96 (70.59) | 7301 (66.79) |
| **Race, n (%) — More than one race** | 3821 | 0 (0.00) | 0 (0.00) | 0 (0.00) | 0 (0.00) | 0 (0.00) | 0 (0.00) |
| **Hispanic or Latino, n (%)** | 429 | 3068 (17.59) | 123 (10.03) | 295 (14.45) | 36 (15.58) | 33 (19.30) | 2581 (18.74) |
| **sPESI score, median [Q1,Q3]** | 0 | 1.0 [0.0,3.0] | 0.0 [0.0,0.0] | 2.0 [1.0,3.0] | 3.0 [2.0,3.0] | 3.0 [3.0,4.0] | 1.0 [1.0,3.0] |
| **PESI score, median [Q1,Q3]** | 0 | 85.7 [61.1,116.0] | 54.4 [39.3,66.0] | 99.6 [82.2,121.3] | 131.6 [108.3,154.4] | 139.8 [117.0,161.8] | 86.6 [61.6,116.3] |
| **Bova score, median [Q1,Q3]** | 0 | 1.0 [0.0,2.0] | 0.0 [0.0,2.0] | 2.0 [1.0,3.0] | 2.0 [1.0,3.0] | 2.0 [1.0,3.0] | 1.0 [0.0,2.0] |
| **Charlson index, median [Q1,Q3]** | 0 | 1.0 [0.0,4.0] | 0.0 [0.0,0.0] | 1.0 [0.0,5.0] | 2.0 [0.0,6.0] | 3.0 [1.0,6.0] | 2.0 [0.0,4.0] |
| **Cancer, n (%)** |  | 5306 (29.69) | 0 (0.00) | 713 (34.13) | 80 (33.61) | 61 (34.46) | 4452 (31.80) |
| **Cardiopulmonary disease, n (%)** |  | 6011 (33.64) | 0 (0.00) | 644 (30.83) | 79 (33.19) | 98 (55.37) | 5190 (37.07) |
| **Heart failure, n (%)** | 17869 |  |  |  |  |  |  |
| **Chronic lung disease, n (%)** | 17869 |  |  |  |  |  |  |
| **Heart rate (max), median [Q1,Q3]** | 1973 | 104.0 [89.0,122.0] | 91.0 [82.0,99.5] | 112.0 [97.0,126.0] | 121.0 [105.0,139.0] | 128.0 [112.0,146.0] | 104.0 [88.0,122.0] |
| **SBP (min), median [Q1,Q3]** | 1975 | 107.0 [96.0,120.0] | 115.0 [107.0,124.0] | 105.0 [96.0,116.0] | 77.0 [70.0,83.0] | 70.0 [61.0,80.0] | 108.0 [96.0,120.0] |
| **MAP (min), median [Q1,Q3]** | 2254 | 79.0 [71.0,88.0] | 84.0 [78.0,92.0] | 78.0 [71.0,86.0] | 58.0 [52.0,63.0] | 57.0 [48.0,67.0] | 80.0 [71.0,89.0] |
| **Resp rate (max), median [Q1,Q3]** | 2021 | 22.0 [20.0,28.0] | 20.0 [18.0,22.0] | 24.0 [20.0,28.0] | 28.0 [23.0,34.0] | 33.0 [28.0,39.0] | 22.0 [20.0,28.0] |
| **SpO2 (min), median [Q1,Q3]** | 1990 | 94.0 [90.0,96.0] | 95.0 [93.0,96.0] | 92.0 [89.0,95.0] | 90.0 [86.0,93.0] | 89.0 [80.0,93.0] | 94.0 [91.0,96.0] |
| **Temp (min), median [Q1,Q3]** | 2031 | 36.5 [36.4,36.7] | 36.5 [36.4,36.7] | 36.5 [36.3,36.6] | 36.4 [36.2,36.6] | 36.3 [35.8,36.6] | 36.5 [36.4,36.7] |
| **Troponin positive, n (%)** |  | 2778 (15.55) | 244 (17.86) | 529 (25.32) | 82 (34.45) | 63 (35.59) | 1860 (13.29) |
| **BNP positive, n (%)** |  | 2223 (12.44) | 20 (1.46) | 326 (15.61) | 55 (23.11) | 41 (23.16) | 1781 (12.72) |
| **Lactate >2 mmol/L, n (%)** |  | 1238 (6.93) | 4 (0.29) | 98 (4.69) | 54 (22.69) | 80 (45.20) | 1002 (7.16) |
| **Creatinine rise, n (%)** |  | 927 (5.19) | 54 (3.95) | 94 (4.50) | 33 (13.87) | 54 (30.51) | 692 (4.94) |
| **RV dysfunction on CT, n (%)** |  | 1107 (6.20) | 155 (11.35) | 411 (19.67) | 59 (24.79) | 56 (31.64) | 426 (3.04) |
| **RV dysfunction on TTE, n (%)** | 17869 |  |  |  |  |  |  |
| **Subsegmental-only clot, n (%)** |  | 290 (1.62) | 93 (6.81) | 172 (8.23) | 20 (8.40) | 5 (2.82) | 0 (0.00) |
| **Hemodynamic instability, n (%)** |  | 853 (4.77) | 0 (0.00) | 0 (0.00) | 0 (0.00) | 156 (88.14) | 697 (4.98) |
| **Vasopressor, n (%)** |  | 748 (4.19) | 0 (0.00) | 0 (0.00) | 0 (0.00) | 142 (80.23) | 606 (4.33) |
| **Inotrope, n (%)** |  | 144 (0.81) | 0 (0.00) | 0 (0.00) | 0 (0.00) | 25 (14.12) | 119 (0.85) |
| **ECMO, n (%)** |  | 39 (0.22) | 0 (0.00) | 0 (0.00) | 0 (0.00) | 22 (12.43) | 17 (0.12) |
| **Cardiac arrest, n (%)** |  | 184 (1.03) | 0 (0.00) | 0 (0.00) | 0 (0.00) | 34 (19.21) | 150 (1.07) |
| **ESC 2019 stratum, n (%) — Low** |  | 1015 (26.23) | 1009 (73.87) | 4 (0.19) | 2 (0.84) | 0 (0.00) | — |
| **ESC 2019 stratum, n (%) — Intermediate-Low** |  | 2332 (60.26) | 357 (26.13) | 1915 (91.67) | 59 (24.79) | 1 (0.56) | — |
| **ESC 2019 stratum, n (%) — Intermediate-High** |  | 171 (4.42) | 0 (0.00) | 170 (8.14) | 1 (0.42) | 0 (0.00) | — |
| **ESC 2019 stratum, n (%) — High** |  | 352 (9.10) | 0 (0.00) | 0 (0.00) | 176 (73.95) | 176 (99.44) | — |
| **Hospital LOS, days, median [Q1,Q3]** | 2262 | 2.4 [0.3,7.2] | 1.8 [0.7,5.7] | 4.9 [2.2,10.7] | 9.4 [4.0,21.3] | 13.9 [6.0,28.4] | 1.9 [0.2,6.3] |
| **In-hospital mortality, n (%)** | 17869 |  |  |  |  |  |  |
| **30-day mortality, n (%)** |  | 1020 (5.71) | 21 (1.54) | 140 (6.70) | 46 (19.33) | 46 (25.99) | 767 (5.48) |
| **1-year mortality, n (%)** |  | 2388 (13.36) | 66 (4.83) | 340 (16.28) | 69 (28.99) | 56 (31.64) | 1857 (13.27) |

##### Table S1c. MIMIC-IV Cohort

| **Characteristic** | **Missing** | **Overall** | **B** | **C** | **D** | **E** | **PE-** |
| --- | --- | --- | --- | --- | --- | --- | --- |
| **n** |  | 4902 | 100 | 214 | 21 | 26 | 4541 |
| **Age, years, median [Q1,Q3]** | 0 | 65.0 [53.0,74.8] | 59.0 [39.8,68.2] | 65.0 [51.0,76.8] | 61.0 [52.0,73.0] | 65.0 [56.5,80.2] | 65.0 [53.0,75.0] |
| **Female sex, n (%)** |  | 2748 (56.06) | 57 (57.00) | 132 (61.68) | 15 (71.43) | 14 (53.85) | 2530 (55.71) |
| **Race, n (%) — American Indian or Alaska Native** | 642 | 5 (0.12) | 0 (0.00) | 0 (0.00) | 0 (0.00) | 0 (0.00) | 5 (0.13) |
| **Race, n (%) — Asian** |  | 190 (4.46) | 3 (3.16) | 9 (4.81) | 0 (0.00) | 0 (0.00) | 178 (4.52) |
| **Race, n (%) — Black or African American** |  | 887 (20.82) | 23 (24.21) | 38 (20.32) | 3 (15.79) | 3 (15.00) | 820 (20.82) |
| **Race, n (%) — Native Hawaiian or Other Pacific Islander** |  | 5 (0.12) | 0 (0.00) | 0 (0.00) | 0 (0.00) | 0 (0.00) | 5 (0.13) |
| **Race, n (%) — White** |  | 3172 (74.46) | 69 (72.63) | 140 (74.87) | 16 (84.21) | 17 (85.00) | 2930 (74.38) |
| **Race, n (%) — More than one race** |  | 1 (0.02) | 0 (0.00) | 0 (0.00) | 0 (0.00) | 0 (0.00) | 1 (0.03) |
| **Hispanic or Latino, n (%)** | 324 | 296 (6.47) | 2 (2.06) | 9 (4.57) | 0 (0.00) | 0 (0.00) | 285 (6.71) |
| **sPESI score, median [Q1,Q3]** | 0 | 1.0 [1.0,2.0] | 0.0 [0.0,0.0] | 1.0 [1.0,2.0] | 3.0 [2.0,4.0] | 3.5 [3.0,4.8] | 1.0 [1.0,2.0] |
| **PESI score, median [Q1,Q3]** | 0 | 92.0 [70.0,117.0] | 59.0 [39.8,69.0] | 94.5 [82.0,115.8] | 146.0 [128.0,167.0] | 146.5 [113.8,176.8] | 93.0 [70.0,117.0] |
| **Bova score, median [Q1,Q3]** | 0 | 0.0 [0.0,2.0] | 0.0 [0.0,2.0] | 1.0 [0.0,2.0] | 2.0 [1.0,3.0] | 3.0 [2.2,4.8] | 0.0 [0.0,2.0] |
| **Charlson index, median [Q1,Q3]** | 0 | 2.0 [1.0,4.0] | 0.0 [0.0,1.0] | 2.0 [1.0,5.0] | 3.0 [1.0,6.0] | 3.0 [2.0,5.8] | 2.0 [1.0,4.0] |
| **Cancer, n (%)** |  | 1396 (28.48) | 0 (0.00) | 92 (42.99) | 10 (47.62) | 9 (34.62) | 1285 (28.30) |
| **Cardiopulmonary disease, n (%)** |  | 2654 (54.14) | 0 (0.00) | 111 (51.87) | 6 (28.57) | 19 (73.08) | 2518 (55.45) |
| **Heart failure, n (%)** |  | 1202 (24.52) | 0 (0.00) | 44 (20.56) | 0 (0.00) | 11 (42.31) | 1147 (25.26) |
| **Chronic lung disease, n (%)** |  | 2182 (44.51) | 0 (0.00) | 94 (43.93) | 6 (28.57) | 15 (57.69) | 2067 (45.52) |
| **Heart rate (max), median [Q1,Q3]** | 1644 | 100.0 [86.0,118.0] | 86.0 [77.5,97.5] | 100.5 [86.0,120.0] | 122.0 [101.0,133.0] | 123.0 [113.0,137.0] | 100.0 [86.0,118.0] |
| **SBP (min), median [Q1,Q3]** | 1648 | 106.0 [94.0,120.0] | 117.0 [111.0,126.0] | 110.0 [98.0,124.0] | 81.0 [79.0,85.0] | 83.0 [76.0,84.5] | 106.0 [94.0,120.0] |
| **MAP (min), median [Q1,Q3]** | 3862 | 61.5 [54.0,70.2] | 80.0 [78.0,84.5] | 74.0 [69.0,80.5] | 55.0 [51.8,57.0] | 55.5 [52.0,63.0] | 61.0 [54.0,70.0] |
| **Resp rate (max), median [Q1,Q3]** | 1645 | 22.0 [20.0,28.0] | 20.0 [18.0,23.5] | 22.0 [18.0,27.0] | 29.0 [26.0,31.0] | 28.0 [27.0,33.0] | 22.0 [20.0,28.0] |
| **SpO2 (min), median [Q1,Q3]** | 1647 | 94.0 [91.0,96.0] | 96.0 [94.5,98.0] | 94.0 [92.0,96.8] | 92.0 [89.0,93.0] | 90.0 [86.0,92.0] | 94.0 [91.0,96.0] |
| **Temp (min), median [Q1,Q3]** | 1690 | 36.6 [36.3,36.7] | 36.7 [36.5,36.8] | 36.6 [36.2,36.8] | 36.3 [36.2,36.6] | 36.5 [36.1,36.9] | 36.6 [36.3,36.7] |
| **Troponin positive, n (%)** |  | 593 (12.10) | 8 (8.00) | 28 (13.08) | 4 (19.05) | 12 (46.15) | 541 (11.91) |
| **BNP positive, n (%)** |  | 194 (3.96) | 2 (2.00) | 21 (9.81) | 1 (4.76) | 2 (7.69) | 168 (3.70) |
| **Lactate >2 mmol/L, n (%)** |  | 416 (8.49) | 1 (1.00) | 13 (6.07) | 3 (14.29) | 7 (26.92) | 392 (8.63) |
| **Creatinine rise, n (%)** |  | 246 (5.02) | 0 (0.00) | 8 (3.74) | 0 (0.00) | 5 (19.23) | 233 (5.13) |
| **RV dysfunction on CT, n (%)** |  | 113 (2.31) | 18 (18.00) | 30 (14.02) | 2 (9.52) | 9 (34.62) | 54 (1.19) |
| **RV dysfunction on TTE, n (%)** |  | 653 (13.32) | 14 (14.00) | 44 (20.56) | 6 (28.57) | 14 (53.85) | 575 (12.66) |
| **Subsegmental-only clot, n (%)** |  | 74 (1.51) | 12 (12.00) | 44 (20.56) | 6 (28.57) | 4 (15.38) | 8 (0.18) |
| **Hemodynamic instability, n (%)** |  | 279 (5.69) | 0 (0.00) | 0 (0.00) | 0 (0.00) | 25 (96.15) | 254 (5.59) |
| **Vasopressor, n (%)** |  | 277 (5.65) | 0 (0.00) | 0 (0.00) | 0 (0.00) | 25 (96.15) | 252 (5.55) |
| **Inotrope, n (%)** |  | 16 (0.33) | 0 (0.00) | 0 (0.00) | 0 (0.00) | 2 (7.69) | 14 (0.31) |
| **ECMO, n (%)** |  | 2 (0.04) | 0 (0.00) | 0 (0.00) | 0 (0.00) | 1 (3.85) | 1 (0.02) |
| **Cardiac arrest, n (%)** |  | 71 (1.45) | 0 (0.00) | 0 (0.00) | 0 (0.00) | 1 (3.85) | 70 (1.54) |
| **ESC 2019 stratum, n (%) — Low** |  | 70 (19.39) | 70 (70.00) | 0 (0.00) | 0 (0.00) | 0 (0.00) | — |
| **ESC 2019 stratum, n (%) — Intermediate-Low** |  | 239 (66.20) | 30 (30.00) | 196 (91.59) | 13 (61.90) | 0 (0.00) | — |
| **ESC 2019 stratum, n (%) — Intermediate-High** |  | 18 (4.99) | 0 (0.00) | 18 (8.41) | 0 (0.00) | 0 (0.00) | — |
| **ESC 2019 stratum, n (%) — High** |  | 34 (9.42) | 0 (0.00) | 0 (0.00) | 8 (38.10) | 26 (100.00) | — |
| **Hospital LOS, days, median [Q1,Q3]** | 0 | 4.4 [1.8,9.3] | 2.5 [1.4,5.0] | 4.9 [2.6,12.0] | 8.7 [4.9,18.0] | 10.7 [6.9,17.1] | 4.3 [1.8,9.2] |
| **In-hospital mortality, n (%)** |  | 225 (4.59) | 0 (0.00) | 8 (3.74) | 1 (4.76) | 7 (26.92) | 209 (4.60) |
| **30-day mortality, n (%)** |  | 378 (7.71) | 0 (0.00) | 18 (8.41) | 2 (9.52) | 8 (30.77) | 350 (7.71) |
| **1-year mortality, n (%)** |  | 1148 (23.42) | 0 (0.00) | 62 (28.97) | 7 (33.33) | 11 (42.31) | 1068 (23.52) |

#### Supplemental Table S2. Mortality by AHA/ACC clinical category and subcategory at 30 days, in hospital, and 1 year.

For each AHA/ACC category (B through E) and subcategory (B1 through E2), the number of patients, number of deaths, mortality (%), and 95% CI are reported. Subcategory estimates are provided for the Duke and INSPECT cohorts; MIMIC-IV is reported at the category level and is denoted imprecise at the subcategory level because of small cell counts. ACC indicates American College of Cardiology; AHA, American Heart Association; CI, confidence interval; INSPECT, Integrating Numerous Sources for Prognostic Evaluation of Clinical Timelines; MIMIC-IV, Medical Information Mart for Intensive Care IV; PE, pulmonary embolism.

| **Cohort** | **Level** | **Group** | **N** | **30-d mortality, n (%) [95% CI]** | **1-y mortality, n (%) [95% CI]** | **In-hosp mortality, n (%) [95% CI]** |
| --- | --- | --- | --- | --- | --- | --- |
| duke | category | B | 2,888 | 46 (1.6%) [1.2–2.1] | 200 (6.9%) [6.1–7.9] | 19 (0.7%) [0.4–1.0] |
| duke | category | C | 7,633 | 726 (9.5%) [8.9–10.2] | 2,010 (26.3%) [25.4–27.3] | 251 (3.3%) [2.9–3.7] |
| duke | category | D | 1,115 | 165 (14.8%) [12.8–17.0] | 380 (34.1%) [31.4–36.9] | 77 (6.9%) [5.6–8.5] |
| duke | category | E | 1,356 | 443 (32.7%) [30.2–35.2] | 633 (46.7%) [44.0–49.3] | 386 (28.5%) [26.1–30.9] |
| duke | subcategory | B1 | 136 | 0 (0.0%) [0.0–2.7] | 11 (8.1%) [4.6–13.9] | 0 (0.0%) [0.0–2.7] |
| duke | subcategory | B2 | 2,752 | 46 (1.7%) [1.3–2.2] | 189 (6.9%) [6.0–7.9] | 19 (0.7%) [0.4–1.1] |
| duke | subcategory | C1 | 2,380 | 219 (9.2%) [8.1–10.4] | 615 (25.8%) [24.1–27.6] | 60 (2.5%) [2.0–3.2] |
| duke | subcategory | C2 | 3,015 | 326 (10.8%) [9.8–12.0] | 849 (28.2%) [26.6–29.8] | 115 (3.8%) [3.2–4.6] |
| duke | subcategory | C3 | 2,238 | 181 (8.1%) [7.0–9.3] | 546 (24.4%) [22.7–26.2] | 76 (3.4%) [2.7–4.2] |
| duke | subcategory | D1 | 783 | 85 (10.9%) [8.9–13.2] | 231 (29.5%) [26.4–32.8] | 40 (5.1%) [3.8–6.9] |
| duke | subcategory | D2 | 332 | 80 (24.1%) [19.8–29.0] | 149 (44.9%) [39.6–50.3] | 37 (11.1%) [8.2–15.0] |
| duke | subcategory | E1 | 1,208 | 375 (31.0%) [28.5–33.7] | 551 (45.6%) [42.8–48.4] | 321 (26.6%) [24.2–29.1] |
| duke | subcategory | E2 | 148 | 68 (45.9%) [38.1–54.0] | 82 (55.4%) [47.4–63.2] | 65 (43.9%) [36.2–52.0] |
| inspect | category | B | 1,366 | 21 (1.5%) [1.0–2.3] | 66 (4.8%) [3.8–6.1] | n/a |
| inspect | category | C | 2,089 | 140 (6.7%) [5.7–7.9] | 340 (16.3%) [14.8–17.9] | n/a |
| inspect | category | D | 238 | 46 (19.3%) [14.8–24.8] | 69 (29.0%) [23.6–35.1] | n/a |
| inspect | category | E | 177 | 46 (26.0%) [20.1–32.9] | 56 (31.6%) [25.2–38.8] | n/a |
| inspect | subcategory | B1 | 93 | 2 (2.2%) [0.6–7.5] | 8 (8.6%) [4.4–16.1] | n/a |
| inspect | subcategory | B2 | 1,273 | 19 (1.5%) [1.0–2.3] | 58 (4.6%) [3.5–5.8] | n/a |
| inspect | subcategory | C1 | 1,201 | 66 (5.5%) [4.3–6.9] | 195 (16.2%) [14.3–18.4] | n/a |
| inspect | subcategory | C2 | 650 | 61 (9.4%) [7.4–11.9] | 116 (17.8%) [15.1–21.0] | n/a |
| inspect | subcategory | C3 | 238 | 13 (5.5%) [3.2–9.1] | 29 (12.2%) [8.6–17.0] | n/a |
| inspect | subcategory | D1 | 160 | 21 (13.1%) [8.7–19.2] | 37 (23.1%) [17.3–30.2] | n/a |
| inspect | subcategory | D2 | 78 | 25 (32.1%) [22.7–43.0] | 32 (41.0%) [30.8–52.1] | n/a |
| inspect | subcategory | E1 | 128 | 31 (24.2%) [17.6–32.3] | 40 (31.2%) [23.9–39.7] | n/a |
| inspect | subcategory | E2 | 49 | 15 (30.6%) [19.5–44.5] | 16 (32.7%) [21.2–46.6] | n/a |
| mimic | category | B | 100 | 0 (0.0%) [0.0–3.7] | 0 (0.0%) [0.0–3.7] | 0 (0.0%) [0.0–3.7] |
| mimic | category | C | 214 | 18 (8.4%) [5.4–12.9] | 62 (29.0%) [23.3–35.4] | 8 (3.7%) [1.9–7.2] |
| mimic | category | D | 21 | 2 (9.5%) [2.7–28.9] | 7 (33.3%) [17.2–54.6] | 1 (4.8%) [0.8–22.7] |
| mimic | category | E | 26 | 8 (30.8%) [16.5–50.0] | 11 (42.3%) [25.5–61.1] | 7 (26.9%) [13.7–46.1] |
| mimic | subcategory | B1 | 12 | 0 (0.0%) [0.0–24.2] | 0 (0.0%) [0.0–24.2] | 0 (0.0%) [0.0–24.2] |
| mimic | subcategory | B2 | 88 | 0 (0.0%) [0.0–4.2] | 0 (0.0%) [0.0–4.2] | 0 (0.0%) [0.0–4.2] |
| mimic | subcategory | C1 | 137 | 13 (9.5%) [5.6–15.6] | 43 (31.4%) [24.2–39.6] | 5 (3.6%) [1.6–8.3] |
| mimic | subcategory | C2 | 50 | 2 (4.0%) [1.1–13.5] | 11 (22.0%) [12.8–35.2] | 1 (2.0%) [0.4–10.5] |
| mimic | subcategory | C3 | 27 | 3 (11.1%) [3.9–28.1] | 8 (29.6%) [15.9–48.5] | 2 (7.4%) [2.1–23.4] |
| mimic | subcategory | D1 | 18 | 2 (11.1%) [3.1–32.8] | 5 (27.8%) [12.5–50.9] | 1 (5.6%) [1.0–25.8] |
| mimic | subcategory | D2 | 3 | 0 (0.0%) [0.0–56.1] | 2 (66.7%) [20.8–93.9] | 0 (0.0%) [0.0–56.1] |
| mimic | subcategory | E1 | 24 | 6 (25.0%) [12.0–44.9] | 9 (37.5%) [21.2–57.3] | 5 (20.8%) [9.2–40.5] |
| mimic | subcategory | E2 | 2 | 2 (100.0%) [34.2–100.0] | 2 (100.0%) [34.2–100.0] | 2 (100.0%) [34.2–100.0] |

#### Supplemental Table S3. Distribution of patients across AHA/ACC subcategories.

Counts and column proportions for subcategories B1 through E2 are reported for the Duke, INSPECT, and MIMIC-IV cohorts. ACC indicates American College of Cardiology; AHA, American Heart Association; INSPECT, Integrating Numerous Sources for Prognostic Evaluation of Clinical Timelines; MIMIC-IV, Medical Information Mart for Intensive Care IV.

| **Subcategory** | **Duke** | **INSPECT** | **MIMIC-IV** |
| --- | --- | --- | --- |
| B1 | 136 (1.1%) | 93 (2.4%) | 12 (3.3%) |
| B2 | 2,752 (21.2%) | 1,273 (32.9%) | 88 (24.4%) |
| C1 | 2,380 (18.3%) | 1,201 (31.0%) | 137 (38.0%) |
| C2 | 3,015 (23.2%) | 650 (16.8%) | 50 (13.8%) |
| C3 | 2,238 (17.2%) | 238 (6.2%) | 27 (7.5%) |
| D1 | 783 (6.0%) | 160 (4.1%) | 18 (5.0%) |
| D2 | 332 (2.6%) | 78 (2.0%) | 3 (0.8%) |
| E1 | 1,208 (9.3%) | 128 (3.3%) | 24 (6.7%) |
| E2 | 148 (1.1%) | 49 (1.3%) | 2 (0.6%) |

Supplemental Table S4. Treatment and management by AHA/ACC clinical category and subcategory, Duke cohort. The proportion of patients receiving anticoagulation alone, systemic thrombolysis, catheter-directed thrombolysis, mechanical thrombectomy, interventional radiology procedures, or extracorporeal membrane oxygenation is reported for each stratum. This table is restricted to the Duke cohort, in which advanced-therapy data are available; INSPECT records anticoagulation status only and MIMIC-IV lacks extracted therapy data, so neither is included. ACC indicates American College of Cardiology; AHA, American Heart Association; ECMO, extracorporeal membrane oxygenation; USCDT, ultrasound-assisted catheter-directed thrombolysis.

| **Cohort** | **Level** | **Group** | **N** | **Anticoagulation only** | **Systemic thrombolysis** | **Catheter-directed thrombolysis (non-USCDT)** | **Aspiration/**  **mechanical thrombectomy** | **ECMO** |
| --- | --- | --- | --- | --- | --- | --- | --- | --- |
| duke | category | B | 2,888 | 2,832 (98.1%) | 16 (0.6%) | 8 (0.3%) | 36 (1.2%) | 0 |
| duke | category | C | 7,633 | 7,297 (95.6%) | 115 (1.5%) | 75 (1.0%) | 209 (2.7%) | 0 |
| duke | category | D | 1,115 | 977 (87.6%) | 46 (4.1%) | 30 (2.7%) | 97 (8.7%) | 0 |
| duke | category | E | 1,356 | 1,112 (82.0%) | 134 (9.9%) | 24 (1.8%) | 126 (9.3%) | 83 (6.1%) |
| duke | subcategory | B1 | 136 | 136 (100.0%) | 0 | 0 | 0 | 0 |
| duke | subcategory | B2 | 2,752 | 2,696 (98.0%) | 16 (0.6%) | 8 (0.3%) | 36 (1.3%) | 0 |
| duke | subcategory | C1 | 2,380 | 2,366 (99.4%) | 8 (0.3%) | 1 (0.0%) | 5 (0.2%) | 0 |
| duke | subcategory | C2 | 3,015 | 2,936 (97.4%) | 16 (0.5%) | 12 (0.4%) | 57 (1.9%) | 0 |
| duke | subcategory | C3 | 2,238 | 1,995 (89.1%) | 91 (4.1%) | 62 (2.8%) | 147 (6.6%) | 0 |
| duke | subcategory | D1 | 783 | 701 (89.5%) | 28 (3.6%) | 18 (2.3%) | 56 (7.2%) | 0 |
| duke | subcategory | D2 | 332 | 276 (83.1%) | 18 (5.4%) | 12 (3.6%) | 41 (12.3%) | 0 |
| duke | subcategory | E1 | 1,208 | 1,010 (83.6%) | 112 (9.3%) | 22 (1.8%) | 95 (7.9%) | 0 |
| duke | subcategory | E2 | 148 | 102 (68.9%) | 22 (14.9%) | 2 (1.4%) | 31 (20.9%) | 83 (56.1%) |

#### Supplemental Table S5. Pairwise differences in 30-day mortality within the intermediate band.

Absolute risk differences (percentage points) with 95% Newcombe (method 10) CIs are reported for all pairwise comparisons among subcategories C1, C2, C3, and D1, with indicators for whether each interval lies within the prespecified equivalence margins of ±3.0 and ±2.0 percentage points. Estimates are reported for the Duke and INSPECT cohorts. ACC indicates American College of Cardiology; AHA, American Heart Association; CI, confidence interval; INSPECT, Integrating Numerous Sources for Prognostic Evaluation of Clinical Timelines; pp, percentage points; SE, standard error; TOST, two one-sided tests.

| **Cohort** | **analysis** | **key** | **Difference, pp** | **95% CI** | **SE, pp** | **detail** |
| --- | --- | --- | --- | --- | --- | --- |
| duke | pairwise difference | C1 vs C2 | -1.61 | -3.21 to 0.01 |  | not equivalent |
| duke | pairwise difference | C1 vs C3 | 1.11 | -0.51 to 2.74 |  | within 3pp |
| duke | pairwise difference | C1 vs D1 | -1.65 | -4.27 to 0.69 |  | not equivalent |
| duke | pairwise difference | C2 vs C3 | 2.73 | 1.12 to 4.29 |  | not equivalent |
| duke | pairwise difference | C2 vs D1 | -0.04 | -2.64 to 2.26 |  | within 3pp |
| duke | pairwise difference | C3 vs D1 | -2.77 | -5.37 to -0.44 |  | not equivalent |
| duke | TOST vs pooled band | C1 | -0.61 |  | 0.71 | p_TOST=0.0003; equivalent @3pp |
| duke | TOST vs pooled band | C2 | 1.83 |  | 0.69 | p_TOST=0.0445; equivalent @3pp |
| duke | TOST vs pooled band | C3 | -2.11 |  | 0.69 | p_TOST=0.0995; not equivalent @3pp |
| duke | TOST vs pooled band | D1 | 1.34 |  | 1.16 | p_TOST=0.077; not equivalent @3pp |
| inspect | pairwise difference | C1 vs C2 | -3.89 | -6.63 to -1.42 |  | not equivalent |
| inspect | pairwise difference | C1 vs C3 | 0.03 | -3.80 to 2.70 |  | not equivalent |
| inspect | pairwise difference | C1 vs D1 | -7.63 | -13.84 to -3.02 |  | not equivalent |
| inspect | pairwise difference | C2 vs C3 | 3.92 | -0.25 to 7.27 |  | not equivalent |
| inspect | pairwise difference | C2 vs D1 | -3.74 | -10.17 to 1.29 |  | not equivalent |
| inspect | pairwise difference | C3 vs D1 | -7.66 | -14.17 to -1.96 |  | not equivalent |
| inspect | TOST vs pooled band | C1 | -3.57 |  | 1.1 | p_TOST=0.697; not equivalent @3pp |
| inspect | TOST vs pooled band | C2 | 3.13 |  | 1.29 | p_TOST=0.5402; not equivalent @3pp |
| inspect | TOST vs pooled band | C3 | -1.90 |  | 1.58 | p_TOST=0.2432; not equivalent @3pp |
| inspect | TOST vs pooled band | D1 | 6.42 |  | 2.73 | p_TOST=0.8955; not equivalent @3pp |

#### Supplemental Table S6. Cox proportional-hazards models of 30-day mortality across the intermediate band.

Hazard ratios with 95% CIs (reference, C1), the likelihood-ratio test comparing the four-level subcategory model with a single pooled-band indicator, and the Schoenfeld proportional-hazards test, including the violation observed for C3, are reported for the Duke cohort. Tied event times were handled by the Efron approximation. ACC indicates American College of Cardiology; AHA, American Heart Association; C-index, concordance index; CI, confidence interval; HR, hazard ratio; INSPECT, Integrating Numerous Sources for Prognostic Evaluation of Clinical Timelines; LR, likelihood ratio.

| **Cohort** | **Panel** | **item** | **estimate** | **95% CI** | **p-value** | **Note** |
| --- | --- | --- | --- | --- | --- | --- |
| duke | Cox PH (ref C1) | C1 | 1.00 |  |  | LR p=0.0064; Schoenfeld p=nan |
| duke | Cox PH (ref C1) | C2 | 1.18 | 0.99–1.40 | 0.058 | LR p=0.0064; Schoenfeld p=0.3129 |
| duke | Cox PH (ref C1) | C3 | 0.88 | 0.72–1.07 | 0.192 | LR p=0.0064; Schoenfeld p=0.0002 |
| duke | Cox PH (ref C1) | D1 | 1.19 | 0.93–1.53 | 0.169 | LR p=0.0064; Schoenfeld p=0.5586 |
| duke | Ordered trend | C1->C2->C3->D1 | 0.99 | 0.92–1.07 | 0.877 | Cochran-Armitage z=-0.204, p=0.8382 |
| duke | Harrell C-index | full (B,C1,C2,C3,D1,D2,E1,E2) | 0.69 | 0.683–0.708 |  |  |
| duke | Harrell C-index | collapsed (B,band,D2,E1,E2) | 0.70 | 0.686–0.707 |  |  |
| duke | Harrell C-index | difference (full - collapsed) | -0.00 |  |  |  |
| inspect | Cox PH (ref C1) | C1 | 1.00 |  |  | LR p=0.0003; Schoenfeld p=nan |
| inspect | Cox PH (ref C1) | C2 | 1.75 | 1.23–2.47 | 0.002 | LR p=0.0003; Schoenfeld p=0.5151 |
| inspect | Cox PH (ref C1) | C3 | 0.99 | 0.55–1.79 | 0.970 | LR p=0.0003; Schoenfeld p=0.2881 |
| inspect | Cox PH (ref C1) | D1 | 2.56 | 1.56–4.18 | <0.001 | LR p=0.0003; Schoenfeld p=0.0045 |
| inspect | Ordered trend | C1->C2->C3->D1 | 1.28 | 1.10–1.49 | 0.002 | Cochran-Armitage z=3.121, p=0.0018 |
| inspect | Harrell C-index | full (B,C1,C2,C3,D1,D2,E1,E2) | 0.73 | 0.702–0.760 |  |  |
| inspect | Harrell C-index | collapsed (B,band,D2,E1,E2) | 0.70 | 0.679–0.732 |  |  |
| inspect | Harrell C-index | difference (full - collapsed) | 0.03 |  |  |  |

#### Supplemental Table S7. Thirty-day mortality within the intermediate band among patients treated with anticoagulation alone.

The number of patients, number of deaths, mortality (%), and 95% Wilson CI are reported.. This analysis is one-directional and is confounded by the clinical indications that prompted escalation. AC indicates anticoagulation; ACC, American College of Cardiology; AHA, American Heart Association; CI, confidence interval.

| **Cohort** | **Subcategory** | **N (AC-only)** | **30-d mortality, n (%) [95% CI]** |
| --- | --- | --- | --- |
| duke | C1 | 2,366 | 218 (9.2%) [8.1–10.4] |
| duke | C2 | 2,936 | 324 (11.0%) [10.0–12.2] |
| duke | C3 | 1,995 | 175 (8.8%) [7.6–10.1] |
| duke | D1 | 701 | 83 (11.8%) [9.7–14.4] |

#### Supplemental Table S8. Bounding of the counterfactual untreated 30-day mortality in subcategory C3.

The observed C3 mortality, the escalated fraction, and the lower and upper bounds of untreated mortality are reported, with the upper bound compared against observed D1 mortality, for the Duke cohort. ACC indicates American College of Cardiology; AHA, American Heart Association.

| **Cohort** | **Subcategory** | **N** | **Observed mortality, %** | **Escalated, %** | **Untreated (lower), %** | **Untreated (upper), %** | **Next tier up** | **Next-tier mortality, %** | **Upper exceeds next tier** |
| --- | --- | --- | --- | --- | --- | --- | --- | --- | --- |
| duke | C1 | 2,380 | 9.20 | 0.59 | 9.16 | 9.75 | C2 | 10.81 | No |
| duke | C2 | 3,015 | 10.81 | 2.62 | 10.75 | 13.37 | C3 | 8.09 | Yes |
| duke | C3 | 2,238 | 8.09 | 10.86 | 7.82 | 18.68 | D1 | 10.86 | Yes |
| duke | D1 | 783 | 10.86 | 10.47 | 10.60 | 21.07 | D2 | 24.10 | No |

#### Supplemental Table S9. Sensitivity of intermediate-band 30-day mortality to handling of C-indeterminate patients.

Mortality by subcategory under three handling modes (exclusion, separate group, and reassignment to C1) is reported for the Duke and INSPECT cohorts. ACC indicates American College of Cardiology; AHA, American Heart Association; CI, confidence interval; INSPECT, Integrating Numerous Sources for Prognostic Evaluation of Clinical Timelines.

| **Cohort** | **Mode** | **Group** | **N** | **30-d mortality, n (%) [95% CI]** |
| --- | --- | --- | --- | --- |
| duke | exclusion | C1 | 1,413 | 118 (8.4%) [7.0–9.9] |
| duke | exclusion | C2 | 3,015 | 326 (10.8%) [9.8–12.0] |
| duke | exclusion | C3 | 2,238 | 181 (8.1%) [7.0–9.3] |
| duke | separate | C1 | 1,413 | 118 (8.4%) [7.0–9.9] |
| duke | separate | C2 | 3,015 | 326 (10.8%) [9.8–12.0] |
| duke | separate | C3 | 2,238 | 181 (8.1%) [7.0–9.3] |
| duke | separate | C-indeterminate | 967 | 101 (10.4%) [8.7–12.5] |
| duke | reassign_to_C1 | C1 | 2,380 | 219 (9.2%) [8.1–10.4] |
| duke | reassign_to_C1 | C2 | 3,015 | 326 (10.8%) [9.8–12.0] |
| duke | reassign_to_C1 | C3 | 2,238 | 181 (8.1%) [7.0–9.3] |
| inspect | exclusion | C1 | 558 | 19 (3.4%) [2.2–5.3] |
| inspect | exclusion | C2 | 650 | 61 (9.4%) [7.4–11.9] |
| inspect | exclusion | C3 | 238 | 13 (5.5%) [3.2–9.1] |
| inspect | separate | C1 | 558 | 19 (3.4%) [2.2–5.3] |
| inspect | separate | C2 | 650 | 61 (9.4%) [7.4–11.9] |
| inspect | separate | C3 | 238 | 13 (5.5%) [3.2–9.1] |
| inspect | separate | C-indeterminate | 643 | 47 (7.3%) [5.5–9.6] |
| inspect | reassign_to_C1 | C1 | 1,201 | 66 (5.5%) [4.3–6.9] |
| inspect | reassign_to_C1 | C2 | 650 | 61 (9.4%) [7.4–11.9] |
| inspect | reassign_to_C1 | C3 | 238 | 13 (5.5%) [3.2–9.1] |

#### Supplemental Table S10. Cross-classification of 2019 ESC risk strata and 2026 AHA/ACC clinical categories.

Cell counts and row proportions are reported for the Duke, INSPECT, and MIMIC-IV cohorts. ESC indicates European Society of Cardiology. ACC indicates American College of Cardiology; AHA, American Heart Association; ESC, European Society of Cardiology; INSPECT, Integrating Numerous Sources for Prognostic Evaluation of Clinical Timelines; MIMIC-IV, Medical Information Mart for Intensive Care IV; mort, mortality.

| **Cohort** | **ESC stratum** | **N** | **B** | **C** | **D** | **E** |
| --- | --- | --- | --- | --- | --- | --- |
| duke | Low | 1,206 | 1,142 (94.7%; 2.0% mort) | 51 (4.2%; 5.9% mort) | 12 (1.0%) | 1 (0.1%) |
| duke | Intermediate-Low | 7,956 | 1,746 (21.9%; 1.3% mort) | 5,752 (72.3%; 9.8% mort) | 456 (5.7%; 11.6% mort) | 2 (0.0%) |
| duke | Intermediate-High | 2,026 | 0 | 1,830 (90.3%; 8.6% mort) | 195 (9.6%; 9.7% mort) | 1 (0.0%) |
| duke | High | 1,804 | 0 | 0 | 452 (25.1%; 20.6% mort) | 1,352 (74.9%; 32.8% mort) |
| inspect | Low | 1,015 | 1,009 (99.4%; 1.3% mort) | 4 (0.4%) | 2 (0.2%) | 0 |
| inspect | Intermediate-Low | 2,332 | 357 (15.3%; 2.2% mort) | 1,915 (82.1%; 6.9% mort) | 59 (2.5%; 6.8% mort) | 1 (0.0%) |
| inspect | Intermediate-High | 171 | 0 | 170 (99.4%; 4.7% mort) | 1 (0.6%) | 0 |
| inspect | High | 352 | 0 | 0 | 176 (50.0%; 23.9% mort) | 176 (50.0%; 26.1% mort) |
| mimic | Low | 70 | 70 (100.0%; 0.0% mort) | 0 | 0 | 0 |
| mimic | Intermediate-Low | 239 | 30 (12.6%; 0.0% mort) | 196 (82.0%; 9.2% mort) | 13 (5.4%) | 0 |
| mimic | Intermediate-High | 18 | 0 | 18 (100.0%) | 0 | 0 |
| mimic | High | 34 | 0 | 0 | 8 (23.5%) | 26 (76.5%; 30.8% mort) |

#### Supplemental Table S11. Difference in 30-day mortality between patients reclassified to category D and those remaining in category C, within the ESC intermediate strata.

For the ESC intermediate-low and intermediate-high strata, separately and pooled, mortality in the category D and category C groups and the absolute risk difference (percentage points) with 95% Newcombe (method 10) CIs are reported for the Duke, INSPECT, and MIMIC-IV cohorts. ACC indicates American College of Cardiology; AHA, American Heart Association; CI, confidence interval; ESC, European Society of Cardiology; INSPECT, Integrating Numerous Sources for Prognostic Evaluation of Clinical Timelines; MIMIC-IV, Medical Information Mart for Intensive Care IV; pp, percentage points.

| **Cohort** | **ESC band** | **N (C)** | **N (D)** | **C mortality, n (%)** | **D mortality, n (%)** | **D−C difference, pp [95% CI]** |
| --- | --- | --- | --- | --- | --- | --- |
| duke | Intermediate-Low | 5,752 | 456 | 566 (9.8%) | 53 (11.6%) | 1.8 [-1.0–5.1] |
| duke | Intermediate-High | 1,830 | 195 | 157 (8.6%) | 19 (9.7%) | 1.2 [-2.5–6.3] |
| duke | Intermediate (pooled) | 7,582 | 651 | 723 (9.5%) | 72 (11.1%) | 1.5 [-0.8–4.2] |
| inspect | Intermediate-Low | 1,915 | 59 | 132 (6.9%) | 4 (6.8%) | -0.1 [-4.4–9.3] |
| inspect | Intermediate-High | 170 | 1 | 8 (4.7%) | 0 (0.0%) | -4.7 [-9.0–74.7] |
| inspect | Intermediate (pooled) | 2,085 | 60 | 140 (6.7%) | 4 (6.7%) | -0.0 [-4.3–9.3] |
| mimic | Intermediate-Low | 196 | 13 | 18 (9.2%) | 2 (15.4%) | 6.2 [-5.9–33.3] |
| mimic | Intermediate (pooled) | 214 | 13 | 18 (8.4%) | 2 (15.4%) | 7.0 [-5.0–34.0] |

#### Supplemental Table S12. Cross-classification of 2019 ESC risk strata and 2026 AHA/ACC subcategories.

Cell counts for the mapping of ESC strata to subcategories B1 through E2 are reported for the Duke, INSPECT, and MIMIC-IV cohorts. ACC indicates American College of Cardiology; AHA, American Heart Association; ESC, European Society of Cardiology; INSPECT, Integrating Numerous Sources for Prognostic Evaluation of Clinical Timelines; MIMIC-IV, Medical Information Mart for Intensive Care IV.

| **Cohort** | **ESC stratum** | **B1** | **B2** | **C1** | **C2** | **C3** | **D1** | **D2** | **E1** | **E2** |
| --- | --- | --- | --- | --- | --- | --- | --- | --- | --- | --- |
| duke | Low | 72.00 | 1070.00 | 42.00 | 9.00 | 0.00 | 11.00 | 1.00 | 0.00 | 1.00 |
| duke | Intermediate-Low | 64.00 | 1682.00 | 2338.00 | 3006.00 | 408.00 | 370.00 | 86.00 | 0.00 | 2.00 |
| duke | Intermediate-High | 0.00 | 0.00 | 0.00 | 0.00 | 1830.00 | 130.00 | 65.00 | 0.00 | 1.00 |
| duke | High | 0.00 | 0.00 | 0.00 | 0.00 | 0.00 | 272.00 | 180.00 | 1208.00 | 144.00 |
| inspect | Low | 72.00 | 937.00 | 4.00 | 0.00 | 0.00 | 2.00 | 0.00 | 0.00 | 0.00 |
| inspect | Intermediate-Low | 21.00 | 336.00 | 1197.00 | 650.00 | 68.00 | 49.00 | 10.00 | 0.00 | 1.00 |
| inspect | Intermediate-High | 0.00 | 0.00 | 0.00 | 0.00 | 170.00 | 1.00 | 0.00 | 0.00 | 0.00 |
| inspect | High | 0.00 | 0.00 | 0.00 | 0.00 | 0.00 | 108.00 | 68.00 | 128.00 | 48.00 |
| mimic | Low | 12.00 | 58.00 | 0.00 | 0.00 | 0.00 | 0.00 | 0.00 | 0.00 | 0.00 |
| mimic | Intermediate-Low | 0.00 | 30.00 | 137.00 | 50.00 | 9.00 | 11.00 | 2.00 | 0.00 | 0.00 |
| mimic | Intermediate-High | 0.00 | 0.00 | 0.00 | 0.00 | 18.00 | 0.00 | 0.00 | 0.00 | 0.00 |
| mimic | High | 0.00 | 0.00 | 0.00 | 0.00 | 0.00 | 7.00 | 1.00 | 24.00 | 2.00 |

#### Supplemental Table S13. Availability of risk-stratification variables by cohort.

The availability of each variable used in category assignment, including echocardiography, computed tomographic right ventricular assessment, cardiac biomarkers, and the respiratory (R) modifier, is indicated for the Duke, INSPECT, and MIMIC-IV cohorts. BNP indicates B-type natriuretic peptide; CT, computed tomography; INSPECT, Integrating Numerous Sources for Prognostic Evaluation of Clinical Timelines; MIMIC-IV, Medical Information Mart for Intensive Care IV; NT-proBNP, N-terminal pro–B-type natriuretic peptide; R, respiratory (modifier); RV, right ventricular; RVD, right ventricular dysfunction; TTE, transthoracic echocardiography.

| **Cohort** | **Variable** | **Available** |
| --- | --- | --- |
| duke | Echocardiography (TTE RVD) | Yes |
| duke | CT RV assessment (rvd_ct) | Yes |
| duke | Troponin | Yes |
| duke | BNP/NT-proBNP | Yes |
| duke | Respiratory (R) modifier inputs | Yes |
| inspect | Echocardiography (TTE RVD) | **No** |
| inspect | CT RV assessment (rvd_ct) | Yes |
| inspect | Troponin | Yes |
| inspect | BNP/NT-proBNP | Yes |
| inspect | Respiratory (R) modifier inputs | Yes |
| mimic | Echocardiography (TTE RVD) | Yes |
| mimic | CT RV assessment (rvd_ct) | Yes |
| mimic | Troponin | Yes |
| mimic | BNP/NT-proBNP | Yes |
| mimic | Respiratory (R) modifier inputs | Yes |

#### Supplemental Table S14. PE positivity rates by hospital.

AHA indicates American Heart Association; PE, pulmonary embolism.

| **Hospital** | **Inpatient n** | **PE+ n** | **PE- n** | **PE+ yield** | **30d mort PE+** | **AHA dist (PE+)** |
| --- | --- | --- | --- | --- | --- | --- |
| Hospital A | 49729 | 7773 | 41956 | 15.6% | 966 (12.4%) | B: 1367 \| C: 4571 \| D: 792 \| E: 1043 |
| Hospital B | 21472 | 2325 | 19147 | 10.8% | 187 (8.0%) | B: 594 \| C: 1467 \| D: 162 \| E: 102 |
| Hospital C | 22704 | 2502 | 20202 | 11.0% | 173 (6.9%) | B: 743 \| C: 1460 \| D: 161 \| E: 138 |
| Hospital D | 196 | 15 | 181 | 7.7% | 0 (0.0%) | B: 5 \| C: 10 \| D: 0 \| E: 0 |
| Other / unrecorded | 1659 | 377 | 1282 | 22.7% | 54 (14.3%) | B: 179 \| C: 125 \| D: 0 \| E: 73 |
| TOTAL | 95760 | 12992 | 82768 | 13.6% |  |  |

#### Supplemental Table S15. RV dysfunction by AHA Sub-Category.

This table demonstrates the proportion of patients with RV dysfunction identified by CT, echocardiography, or both. Note that the C-indeterminate group by definition does not display RV dysfunction on CT and also does not have a TTE to demonstrate RV strain and is reported as 0. ACC indicates American College of Cardiology; AHA, American Heart Association; RV, right ventricular.

| **Cohort** | **Measure** | **Overall** | **B1** | **B2** | **C1** | **C-indeterminate** | **C2** | **C3** | **D1** | **D2** | **E1** | **E2** |
| --- | --- | --- | --- | --- | --- | --- | --- | --- | --- | --- | --- | --- |
| duke | n | 12,992 | 136 | 2,752 | 1,413 | 967 | 3,015 | 2,238 | 783 | 332 | 1,208 | 148 |
| duke | RV dysfunction on CT | 4,064 (31.3%) | 10 (7.4%) | 754 (27.4%) | 0 (0.0%) | 0 (0.0%) | 740 (24.5%) | 1,592 (71.1%) | 287 (36.7%) | 148 (44.6%) | 460 (38.1%) | 73 (49.3%) |
| duke | RV strain on TTE | 3,900 (30.0%) | 11 (8.1%) | 526 (19.1%) | 0 (0.0%) | 0 (0.0%) | 687 (22.8%) | 1,608 (71.8%) | 264 (33.7%) | 151 (45.5%) | 584 (48.3%) | 69 (46.6%) |
| duke | RV strain (CT OR TTE) | 5,884 (45.3%) | 19 (14.0%) | 1,028 (37.4%) | 0 (0.0%) | 0 (0.0%) | 1,176 (39.0%) | 2,238 (100.0%) | 400 (51.1%) | 196 (59.0%) | 730 (60.4%) | 97 (65.5%) |
| duke | RV strain (CT AND TTE) | 2,080 (16.0%) | 2 (1.5%) | 252 (9.2%) | 0 (0.0%) | 0 (0.0%) | 251 (8.3%) | 962 (43.0%) | 151 (19.3%) | 103 (31.0%) | 314 (26.0%) | 45 (30.4%) |
| inspect | n | 3,870 | 93 | 1,273 | 558 | 643 | 650 | 238 | 160 | 78 | 128 | 49 |
| inspect | RV dysfunction on CT | 681 (17.6%) | 9 (9.7%) | 146 (11.5%) | 0 (0.0%) | 0 (0.0%) | 173 (26.6%) | 238 (100.0%) | 40 (25.0%) | 19 (24.4%) | 32 (25.0%) | 24 (49.0%) |
| inspect | RV strain on TTE | 0 (0.0%) | 0 (0.0%) | 0 (0.0%) | 0 (0.0%) | 0 (0.0%) | 0 (0.0%) | 0 (0.0%) | 0 (0.0%) | 0 (0.0%) | 0 (0.0%) | 0 (0.0%) |
| inspect | RV strain (CT OR TTE) | 681 (17.6%) | 9 (9.7%) | 146 (11.5%) | 0 (0.0%) | 0 (0.0%) | 173 (26.6%) | 238 (100.0%) | 40 (25.0%) | 19 (24.4%) | 32 (25.0%) | 24 (49.0%) |
| inspect | RV strain (CT AND TTE) | 0 (0.0%) | 0 (0.0%) | 0 (0.0%) | 0 (0.0%) | 0 (0.0%) | 0 (0.0%) | 0 (0.0%) | 0 (0.0%) | 0 (0.0%) | 0 (0.0%) | 0 (0.0%) |
| mimic | n | 361 | 12 | 88 | 52 | 85 | 50 | 27 | 18 | 3 | 24 | 2 |
| mimic | RV dysfunction on CT | 59 (16.3%) | 0 (0.0%) | 18 (20.5%) | 0 (0.0%) | 0 (0.0%) | 16 (32.0%) | 14 (51.9%) | 2 (11.1%) | 0 (0.0%) | 8 (33.3%) | 1 (50.0%) |
| mimic | RV strain on TTE | 78 (21.6%) | 0 (0.0%) | 14 (15.9%) | 0 (0.0%) | 0 (0.0%) | 22 (44.0%) | 22 (81.5%) | 6 (33.3%) | 0 (0.0%) | 13 (54.2%) | 1 (50.0%) |
| mimic | RV strain (CT OR TTE) | 108 (29.9%) | 0 (0.0%) | 26 (29.5%) | 0 (0.0%) | 0 (0.0%) | 32 (64.0%) | 27 (100.0%) | 7 (38.9%) | 0 (0.0%) | 14 (58.3%) | 2 (100.0%) |
| mimic | RV strain (CT AND TTE) | 29 (8.0%) | 0 (0.0%) | 6 (6.8%) | 0 (0.0%) | 0 (0.0%) | 6 (12.0%) | 9 (33.3%) | 1 (5.6%) | 0 (0.0%) | 7 (29.2%) | 0 (0.0%) |

#### Supplemental Table S16. Mortality by advanced therapy.

This table first demonstrates thirty-day mortality by any patient receiving advanced therapy (e.g., aspiration/mechanical thrombectomy, catheter directed thrombolytics, etc.) stratified by AHA/ACC 2026 categories/subcategories, then by ESC to AHA/ACC 2026 subcategories to demonstrate the influence of the reassignment on both intervention and therapy. ACC indicates American College of Cardiology; AHA, American Heart Association; CI, confidence interval; ESC, European Society of Cardiology; PE, pulmonary embolism.

| **Cohort** | **Panel** | **Stratum** | **N** | **Advanced therapy, n (%)** | **30-d mortality — all,**  **n / N (%) [95% CI]** | **30-d mortality —**  **no advanced therapy,**  **n / N (%) [95% CI]** | **30-d mortality —**  **advanced therapy,**  **n / N (%) [95% CI]** |
| --- | --- | --- | --- | --- | --- | --- | --- |
| duke | By AHA/ACC category | B | 2,888 | 56 (1.9%) | 46 / 2,888 (1.6%) [1.2–2.1] | 44 / 2,832 (1.6%) [1.2–2.1] | 2 / 56 (3.6%) [1.0–12.1] |
| duke |  | C | 7,633 | 336 (4.4%) | 726 / 7,633 (9.5%) [8.9–10.2] | 717 / 7,297 (9.8%) [9.2–10.5] | 9 / 336 (2.7%) [1.4–5.0] |
| duke |  | D | 1,115 | 138 (12.4%) | 165 / 1,115 (14.8%) [12.8–17.0] | 159 / 977 (16.3%) [14.1–18.7] | 6 / 138 (4.3%) [2.0–9.2] |
| duke |  | E | 1,356 | 296 (21.8%) | 443 / 1,356 (32.7%) [30.2–35.2] | 351 / 1,060 (33.1%) [30.3–36.0] | 92 / 296 (31.1%) [26.1–36.6] |
| duke |  | B1 | 136 | 0 | 0 / 136 (0.0%) [0.0–2.7] | 0 / 136 (0.0%) [0.0–2.7] | 0 / 0 (–) |
| duke |  | B2 | 2,752 | 56 (2.0%) | 46 / 2,752 (1.7%) [1.3–2.2] | 44 / 2,696 (1.6%) [1.2–2.2] | 2 / 56 (3.6%) [1.0–12.1] |
| duke |  | C1 | 2,380 | 14 (0.6%) | 219 / 2,380 (9.2%) [8.1–10.4] | 218 / 2,366 (9.2%) [8.1–10.4] | 1 / 14 (7.1%) [1.3–31.5] |
| duke |  | C2 | 3,015 | 79 (2.6%) | 326 / 3,015 (10.8%) [9.8–12.0] | 324 / 2,936 (11.0%) [10.0–12.2] | 2 / 79 (2.5%) [0.7–8.8] |
| duke |  | C3 | 2,238 | 243 (10.9%) | 181 / 2,238 (8.1%) [7.0–9.3] | 175 / 1,995 (8.8%) [7.6–10.1] | 6 / 243 (2.5%) [1.1–5.3] |
| duke |  | D1 | 783 | 82 (10.5%) | 85 / 783 (10.9%) [8.9–13.2] | 83 / 701 (11.8%) [9.7–14.4] | 2 / 82 (2.4%) [0.7–8.5] |
| duke |  | D2 | 332 | 56 (16.9%) | 80 / 332 (24.1%) [19.8–29.0] | 76 / 276 (27.5%) [22.6–33.1] | 4 / 56 (7.1%) [2.8–17.0] |
| duke |  | E1 | 1,208 | 198 (16.4%) | 375 / 1,208 (31.0%) [28.5–33.7] | 321 / 1,010 (31.8%) [29.0–34.7] | 54 / 198 (27.3%) [21.5–33.9] |
| duke |  | E2 | 148 | 98 (66.2%) | 68 / 148 (45.9%) [38.1–54.0] | 30 / 50 (60.0%) [46.2–72.4] | 38 / 98 (38.8%) [29.7–48.7] |
| duke | By ESC 2019 → AHA/ACC 2026 | Low → B | 1,142 | 8 (0.7%) | 23 / 1,142 (2.0%) [1.3–3.0] | 22 / 1,134 (1.9%) [1.3–2.9] | 1 / 8 (12.5%) [2.2–47.1] |
| duke |  | Low → C | 51 | 2 (3.9%) | 3 / 51 (5.9%) [2.0–15.9] | 3 / 49 (6.1%) [2.1–16.5] | 0 / 2 (0.0%) [0.0–65.8] |
| duke |  | Low → D | 12 | 0 | 0 / 12 (0.0%) [0.0–24.2] | 0 / 12 (0.0%) [0.0–24.2] | 0 / 0 (–) |
| duke |  | Low → E | 1 | 1 (100.0%) | 0 / 1 (0.0%) [0.0–79.3] | 0 / 0 (–) | 0 / 1 (0.0%) [0.0–79.3] |
| duke |  | Intermediate-Low → B | 1,746 | 48 (2.7%) | 23 / 1,746 (1.3%) [0.9–2.0] | 22 / 1,698 (1.3%) [0.9–2.0] | 1 / 48 (2.1%) [0.4–10.9] |
| duke |  | Intermediate-Low → C | 5,752 | 118 (2.1%) | 566 / 5,752 (9.8%) [9.1–10.6] | 563 / 5,634 (10.0%) [9.2–10.8] | 3 / 118 (2.5%) [0.9–7.2] |
| duke |  | Intermediate-Low → D | 456 | 10 (2.2%) | 53 / 456 (11.6%) [9.0–14.9] | 51 / 446 (11.4%) [8.8–14.7] | 2 / 10 (20.0%) [5.7–51.0] |
| duke |  | Intermediate-Low → E | 2 | 2 (100.0%) | 0 / 2 (0.0%) [0.0–65.8] | 0 / 0 (–) | 0 / 2 (0.0%) [0.0–65.8] |
| duke |  | Intermediate-High → C | 1,830 | 216 (11.8%) | 157 / 1,830 (8.6%) [7.4–10.0] | 151 / 1,614 (9.4%) [8.0–10.9] | 6 / 216 (2.8%) [1.3–5.9] |
| duke |  | Intermediate-High → D | 195 | 71 (36.4%) | 19 / 195 (9.7%) [6.3–14.7] | 18 / 124 (14.5%) [9.4–21.8] | 1 / 71 (1.4%) [0.2–7.6] |
| duke |  | Intermediate-High → E | 1 | 1 (100.0%) | 0 / 1 (0.0%) [0.0–79.3] | 0 / 0 (–) | 0 / 1 (0.0%) [0.0–79.3] |
| duke |  | High → D | 452 | 57 (12.6%) | 93 / 452 (20.6%) [17.1–24.5] | 90 / 395 (22.8%) [18.9–27.2] | 3 / 57 (5.3%) [1.8–14.4] |
| duke |  | High → E | 1,352 | 292 (21.6%) | 443 / 1,352 (32.8%) [30.3–35.3] | 351 / 1,060 (33.1%) [30.3–36.0] | 92 / 292 (31.5%) [26.4–37.0] |

### Supplemental Figures

#### Supplemental Figure S1. Subcategory-level Kaplan-Meier survival.

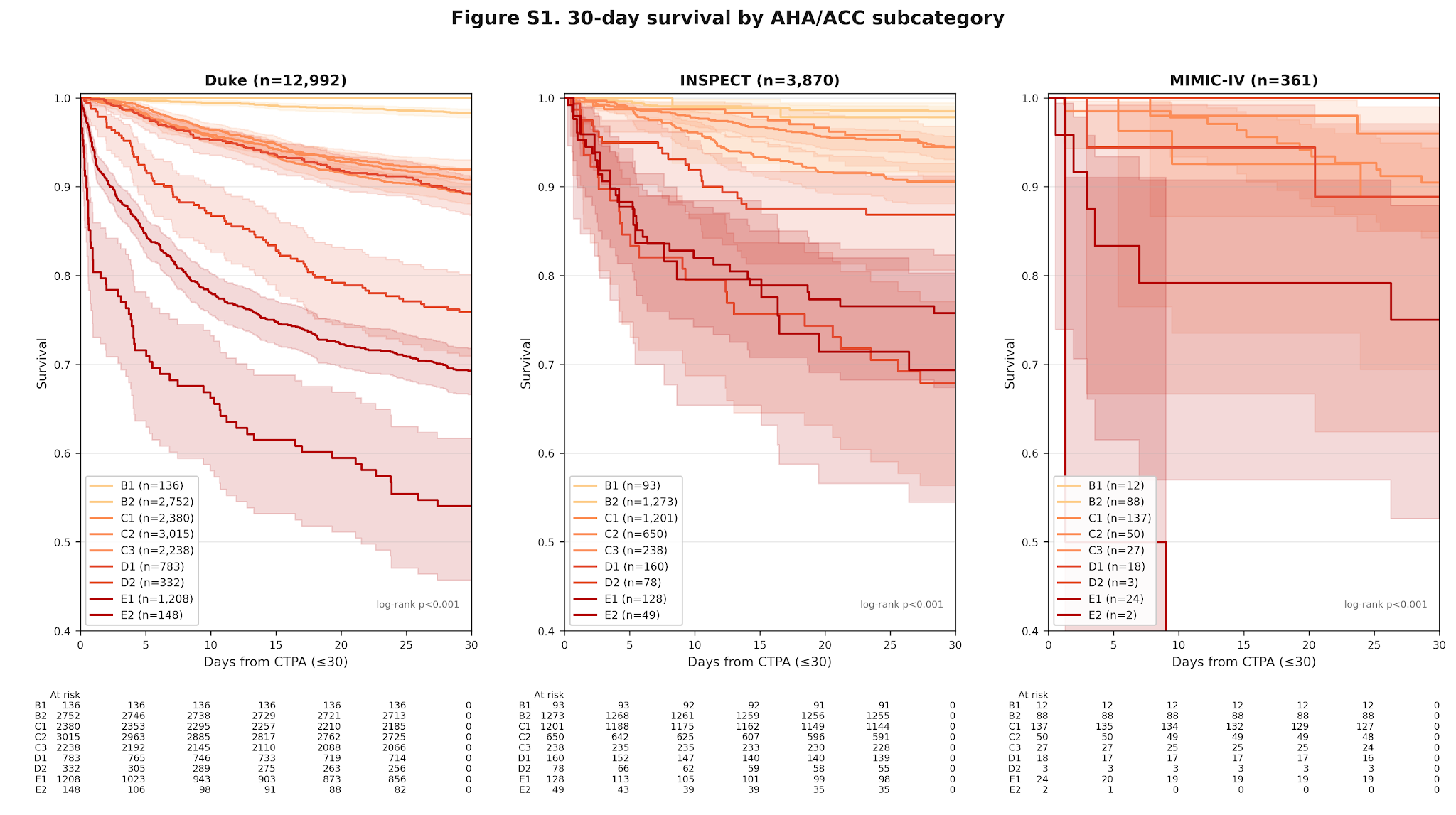

#### Supplemental Figure S2. Forest plots of pairwise differences in 30-day mortality within the intermediate band.

Pairwise risk differences among subcategories C1, C2, C3, and D1, with 95% Newcombe (method 10, score interval for the single proportion) CIs, are shown against the prespecified equivalence margins for the Duke and INSPECT cohorts.
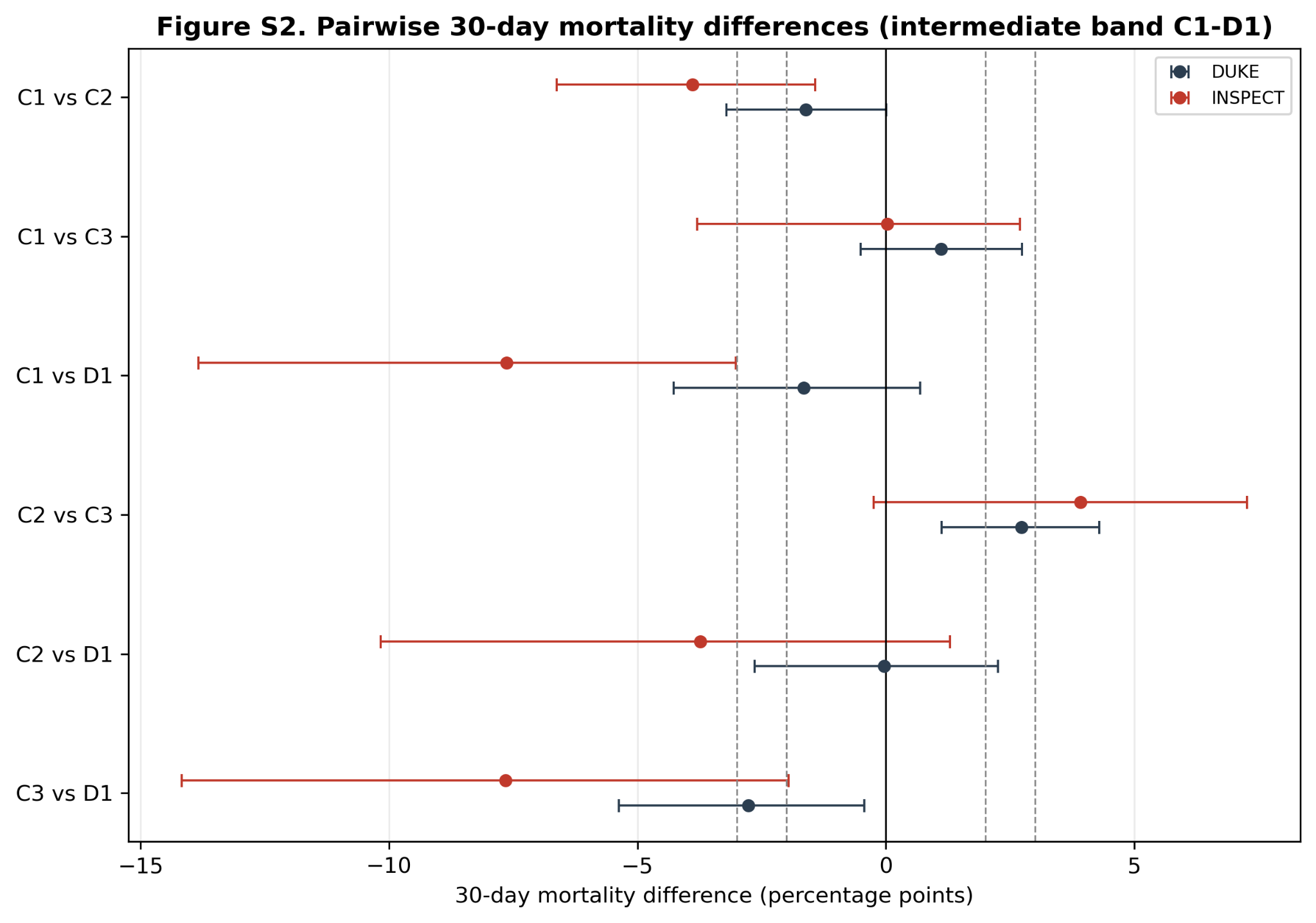

#### Supplemental Figure S3. Reclassification of patients from 2019 ESC risk strata to 2026 AHA/ACC categories, external cohorts.

Alluvial diagrams for the INSPECT and MIMIC-IV cohorts, with the INSPECT diagram interpreted in light of the absence of echocardiography.

##### Fig S3a. INSPECT

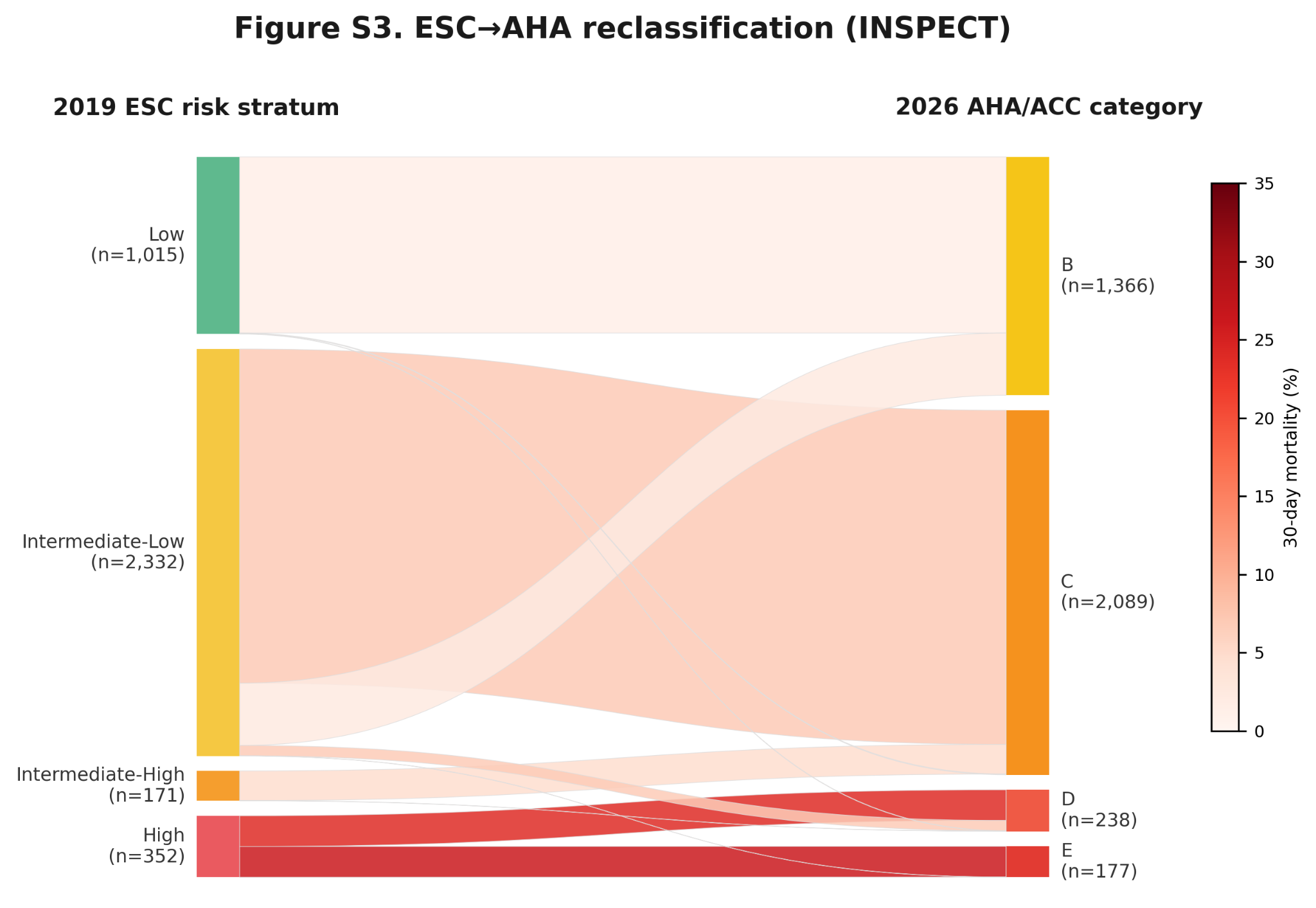

##### Fig S3b. MIMIC-IV

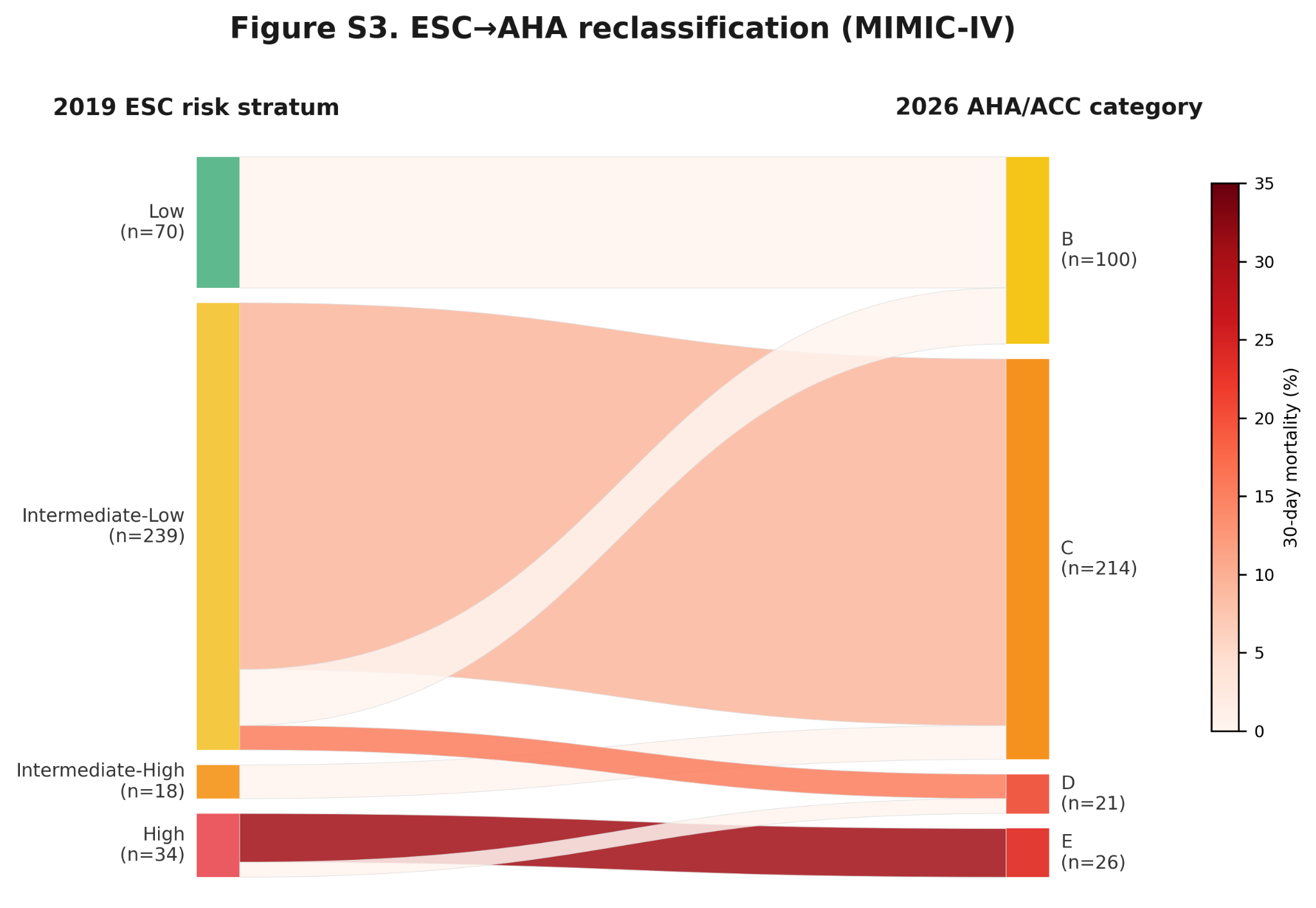

##### Fig S3c. Duke

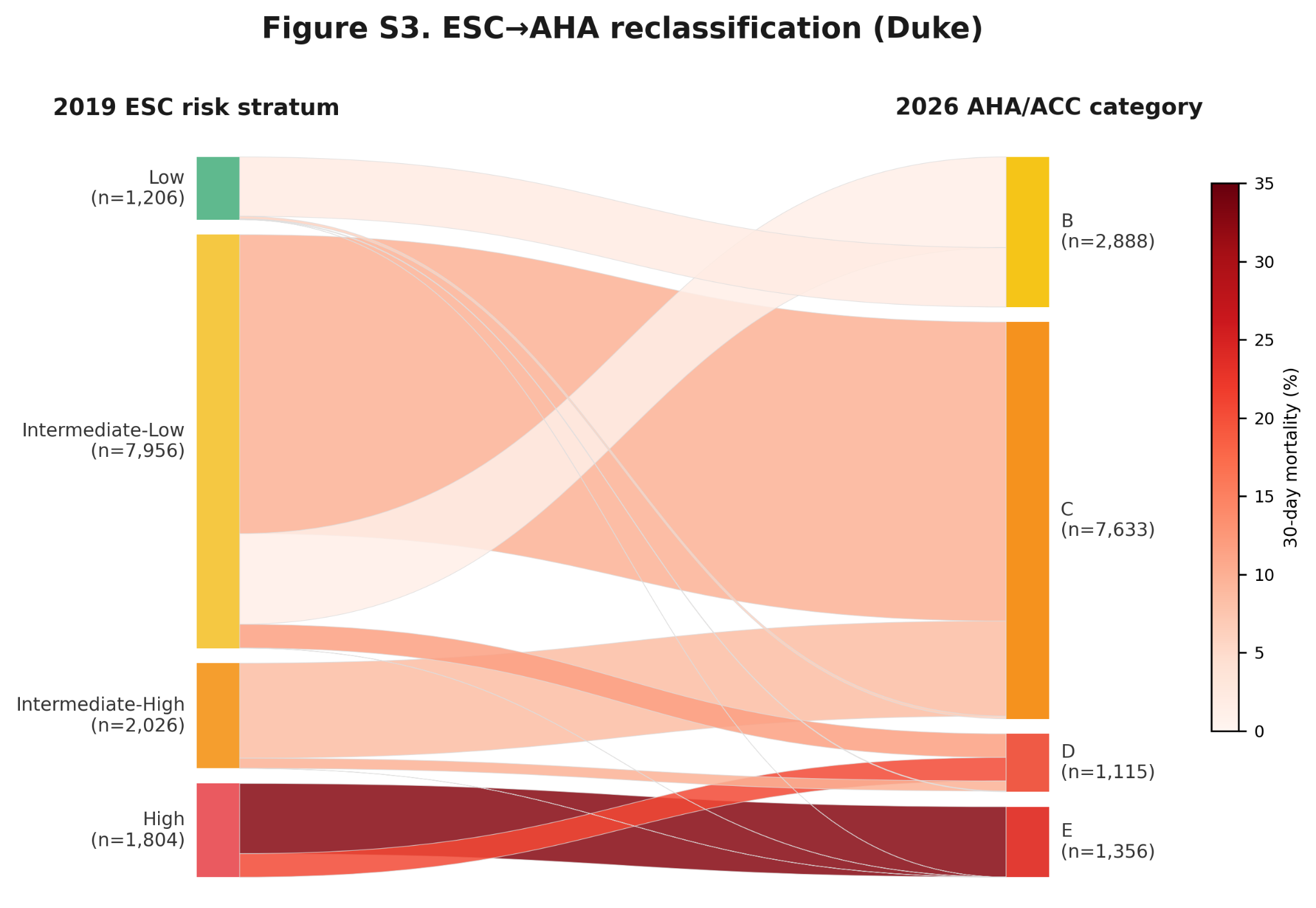

#### Supplemental Figure S4. One-year survival by AHA/ACC clinical category.

Kaplan-Meier survival curves for AHA/ACC categories B through E extended to 1 year.

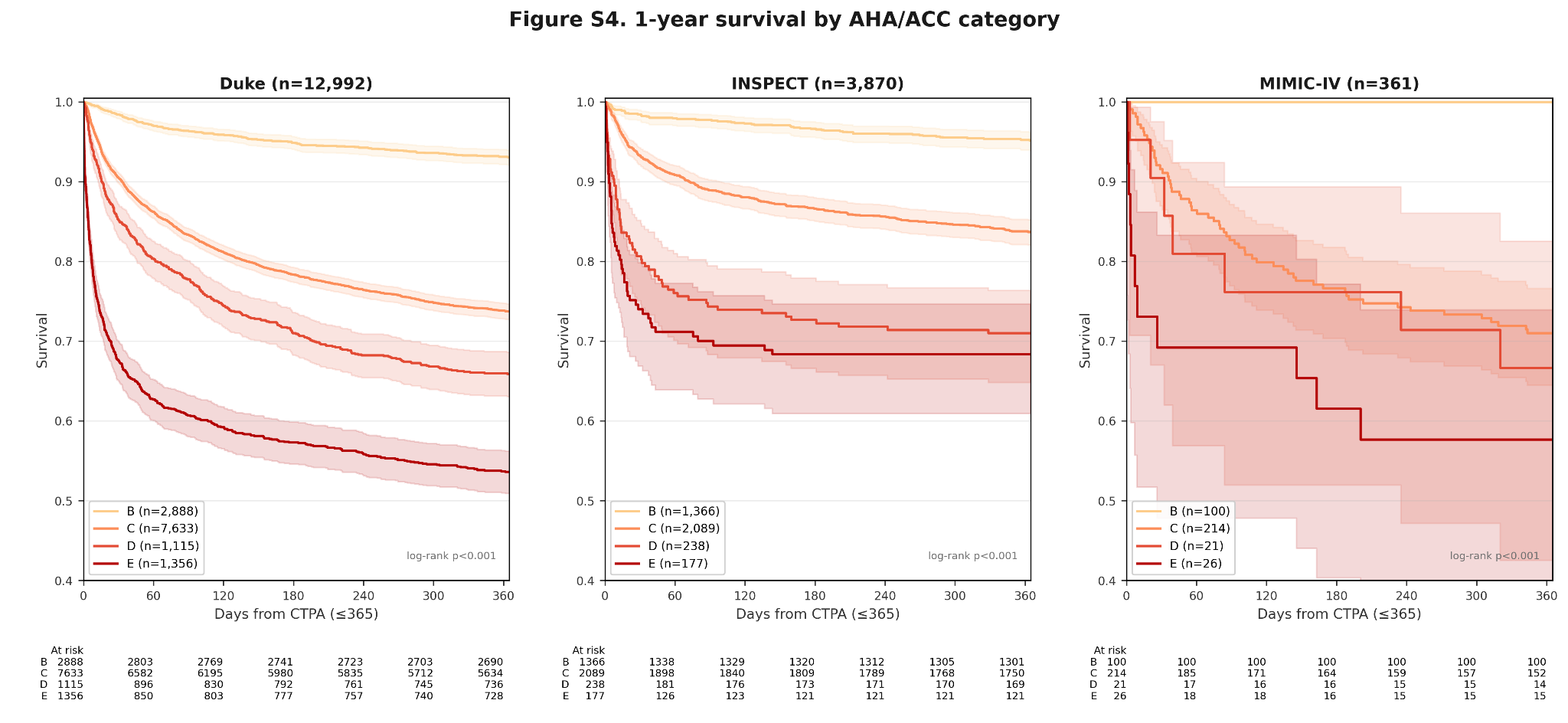
